# Spatiotemporal disease suitability prediction for Oropouche virus and the role of vectors across the Americas

**DOI:** 10.1101/2025.02.28.25323068

**Authors:** Jenicca Poongavanan, Marcel Dunaiski, Graeme Dor, Moritz U.G. Kraemer, Marta Giovanetti, Ahyoung Lim, Oliver J. Brady, Bernardo Gutierrez, Jean Paul Carrera, CLIMADE Consortium, Cheryl Baxter, Vagner Fonseca, Luiz Alcantara, Tulio de Oliveira, Houriiyah Tegally

## Abstract

Oropouche virus (OROV) is an emerging arbovirus with increasing outbreaks in South America, yet its environmental drivers and potential range remain poorly understood. Using ecological niche modeling (ENM) with random forests, we assessed the environmental suitability of OROV and its primary vector, *Culicoides paraensis*, across Brazil and the Americas. We evaluated five pseudo-absence sampling techniques, considering pseudo-absence ratios, buffer radii, and density smoothing factors to determine the most effective modeling approach. Key environmental predictors included humidity, agricultural land-use, and forest cover for both the virus and the vector. The resulting suitability model identifies high transmission risk areas in Central and South America, and reveals that environmental suitability patterns align with seasonal fluctuations in case numbers, with peaks in Amazonian states at the beginning of the year and an expansion into non-Amazonian regions later in the year. A bivariate suitability map highlighted strong spatial overlap between OROV and *Culicoides paraensis*, with potential co-suitability areas with *Culex quinquefasciatus* mosquito, a suspected secondary vector. These findings enhance understanding of OROV transmission dynamics, supporting risk assessment, surveillance, and vector control strategies.

## 1. Introduction

Oropouche virus (OROV) is an emerging arthropod-borne virus responsible for frequent and widespread outbreaks in South America, particularly in Brazil over the last few years. Since its discovery in 1955, OROV has caused numerous epidemics, with over half a million reported cases across several South and Central American countries, including Brazil, Cuba, Peru, Colombia, and Panama ^1,2^. The virus is primarily transmitted by *Culicoides paraensis*, a biting midge that can occur all year round in urban and peri-urban areas^3^ although other potential vectors have been implicated in its transmission cycle such as *Culex quinquefasciatus*^4–7^. The symptoms of Oropouche fever, including high fever, headache, and joint pain, are often misdiagnosed as dengue or other febrile illnesses, contributing to underreporting and gaps in surveillance ^8^. While historically confined to the Amazon Basin and northern Brazil, OROV is increasingly being detected in new geographic regions, raising concerns about its potential expansion^9–11^.

The 2023–2024 OROV outbreak in Brazil has been particularly alarming, with a rapid rise in cases across multiple states and 14,613 reported cases up till early 2025^12^. This surge has been attributed to changing land-use patterns, urbanization, and shifting climate conditions that may alter the ecological niche of both the virus and its vectors^9,11,13^. In 2024, reemergence and rapid epidemic expansion of OROV in Brazil and neighbouring countries^14^, including, for the first time, transmission beyond the Amazon basin^9,10^, and fatal cases^15^. In a previous study, we investigated the link between environmental covariates and OROV transmission in Brazil prior to and following amplified transmission, and pointed to a potential shift in the ecological niche of the virus preceding the epidemic^11^. This underscores a critical need for a comprehensive continental-scale assessment of OROV transmission potential and its vector. Developing detailed suitability maps can provide valuable insights into areas at risk, guiding public health interventions. Incorporating key environmental factors such as climate, land use, and vector ecology will improve our understanding of the conditions that facilitate virus persistence and spread.

Ecological niche modeling (ENM) is widely used in species ecology to predict species distributions (plants and animals) based on environmental conditions. More recently, it has been applied in disease ecology, particularly for vector-borne diseases, to assess transmission risk and identify high-risk areas for outbreaks^16–18^. While ENMs are widely used to predict vector environmental suitability based on presence-only data and environmental variables, their application to enumerate populations at risk of disease has also proved useful, improving surveillance and control strategies^19^. Various ENM approaches exist, including statistical models (e.g., Generalized Linear Models, Generalized Additive Models), envelope models (e.g., Maximum Entropy, Surface Range Envelope), and machine learning algorithms (e.g., Random Forest, Boosted Regression Trees, Artificial Neural Networks). These models often rely on pseudo-absence data when true absence data is unavailable, using these points to contrast presence locations. While random sampling is commonly used to generate pseudo-absence data^20,21^, more informed approaches have been suggested that enhance ecological relevance and potentially improve model accuracy. Informed sampling techniques incorporate spatial or environmental constraints, such as buffer zones around presence points (geographic-weighted exclusion)^22^ or unsuitable environmental conditions (environmental-weighted exclusion)^20,23^. However, the impact of these methods on the resulting model outcomes has not been assessed for OROV.

In this study, we use random forest (RF) modeling to investigate the environmental suitability of OROV and its primary vector, *C. paraensis*. We evaluate the impact of five pseudo-absence sampling techniques to determine the most effective approach for modeling OROV distribution. We generate monthly environmental suitability predictions for OROV across Brazil and spatial continental-scale predictions across the Americas based on the consensus suitability model, which integrates predictions from multiple pseudo-absence sampling techniques. This approach ensures that both seasonal transmission patterns and broader geographic risk areas are captured with improved reliability. Additionally, we examine key environmental predictors influencing OROV transmission and assess the spatial overlap between the virus and its vectors. Finally, we construct bivariate suitability maps to explore potential co-occurrence zones between OROV, *C. paraensis* and *Cx. quinquefasciatus*, the suspected secondary vector. By identifying high-risk areas and key environmental drivers, this study provides critical insights into OROV transmission dynamics, to better inform surveillance and vector control strategies.

## 2. Results

### Performance Across Pseudo-Absence Sampling Strategies

We tested five pseudo-absence sampling strategies under different pseudo-absence ratios: (1) Random sampling, where pseudo-absences were randomly selected across the study area, assuming a uniform probability of species absence; (2) Geographic sampling, which constrained pseudo-absences within a defined radius around presence points (See illustration in **Supplementary S9B)**; (3) Density-weighted geographic sampling, which incorporated the density of presence points into pseudo-absence selection, ensuring absences were more likely to be placed in areas with higher sampling effort, where the virus was likely searched for but not detected; (4) Density-weighted population-based sampling,

which further refined selection by integrating both the density of occurrence points and human population density, emphasizing areas of higher human interaction; and (5) Target group-based sampling, which used the distribution of other arboviruses (dengue, chikungunya, yellow fever, and Zika) to guide pseudo-absence selection, assuming that OROV surveillance would follow a similar spatial pattern.

The performance of ENMs varied across different pseudo-absence (PA) sampling strategies, with notable differences observed across PA ratios, buffer radii, and KDE smoothing factors. To evaluate model performance, we used the Boyce Index (BI), a presence-only metric that assesses how well predicted suitability aligns with independent occurrence data. A higher BI value indicates stronger agreement between the model’s predictions and observed presence locations, making it particularly useful for evaluating models that rely on pseudo-absence data. Among the five sampling techniques assessed, the target group-based Sampling approach yielded the highest BI of 0.96 (ROC = 0.9) under a PA ratio of 1:1, a buffer radius of 50-500 km and a KDE smoothing factor of 5 (**Figure 1A**). In contrast, random sampling consistently produced the lowest BI values, with a maximum of 0.72 at a PA ratio of 10,000 (ROC = 0.509). Model performance was generally better across the different sampling techniques when a PA ratio of 1:1 was applied (See **Supplementary Figure S1**). However, we see less variability in model performance for a balanced presence-absence ratio (1:1, n=400) and for a high pseudo absence number (n=10,000) (See **Supplementary Figure S1**). When examining the effect of buffer radii, we observe less variability in model performances when a more restrictive geographic buffer is applied, in this case between 50 - 150 km and 50 - 300 km (see **Supplementary Figure S1**). Overall, smaller KDE smoothing factors were associated with improved model performance across sampling techniques, PA ratios, and buffer radii, suggesting that less smoothing enhances the model’s ability to capture finer-scale patterns.

**Figure 1.**
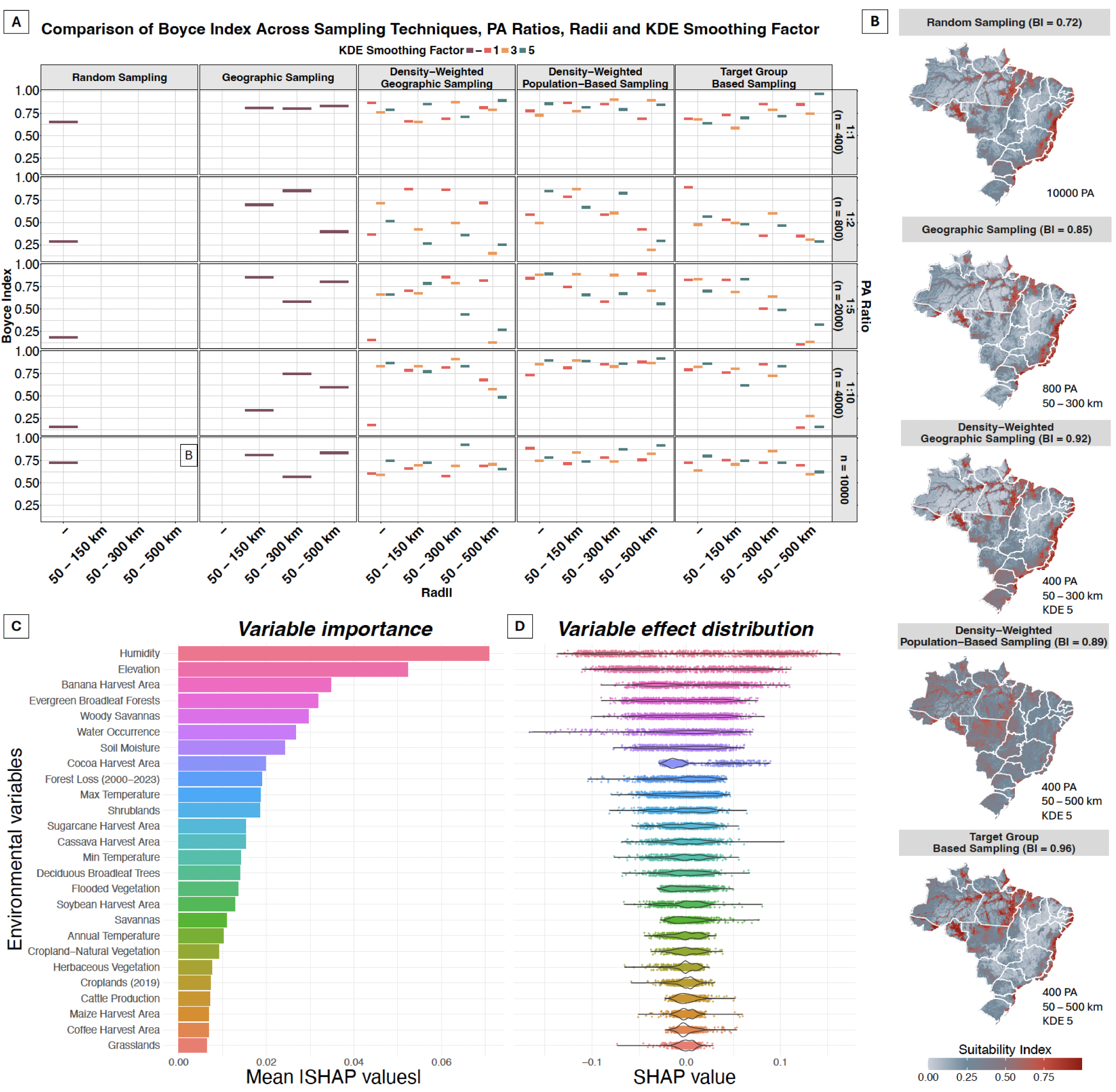
Comparison of sampling techniques, variable importance, and spatial suitability predictions. (A) Boyce Index (BI) performance across five pseudo-absence sampling techniques (Random Sampling, Geographic Sampling, Density-Weighted Geographic Sampling, Density-Weighted Population-Based Sampling, and Target Group-Based Sampling) with varying PA ratios and buffer radii. Higher BI values indicate better model performance. (B) Spatial distribution of habitat suitability predicted under each sampling strategy (predictions from the best performing model under each technique), with darker red areas indicating higher suitability. The BI for each method is displayed in parentheses. (C) Variable importance calculated as the average of absolute SHAP values in the training dataset, indicating the average variable contribution to the overall prediction of the model. (D) shows the distribution of SHAP values per variable, indicating whether contributions are positive or negative.

The spatial predictions of environmental suitability also varied across sampling strategies (**Figure 1B**). Across all methods, regions of highest suitability (red areas) were predominantly concentrated in areas where known virus occurrences have been reported, particularly in the northern and coastal regions of Brazil, including parts of the Amazon Basin. However, the extent and specificity of these high-suitability areas varied depending on the PA sampling technique applied. The best-performing model for geographic sampling produced a more spatially constrained suitability pattern (**Figure 1B**), with high suitability concentrated in regions with reported occurrences, while other areas exhibited very low suitability values. This pattern may be attributed to the restricted buffer of 50–150 km, which likely limited the spatial extent of pseudo-absence selection, resulting in a more localized prediction of environmental suitability. The density-weighted population-based sampling approach generated a broader environmental suitability distribution across the country, while exhibiting a more spatially constrained high-suitability pattern along densely populated coastal regions. This outcome is likely a result of the underlying methodology, in which pseudo-absences were predominantly sampled near populated areas, thereby refining suitability predictions in urbanized regions while maintaining a wider overall suitability range. The target group-based sampling method generated the most spatially precise predictions, with high-suitability areas concentrated along humid, densely forested, and agriculturally active zones.

We used SHAP (Shapley Additive Explanations) values to quantify both the importance and marginal contribution of each variable to predict OROV suitability (**Figure 1 C&D**). The variable importance was computed as the average of absolute SHAP values in the training dataset. We found that humidity was the most influential predictor, contributing 7.08% to the model’s explanatory power (**Figure 1C**). Other key predictors included elevation (5.23%), banana harvested area (3.48%),, and forest cover (3.18%), highlighting the role of land-use and topographic features in shaping environmental suitability for virus transmission. In contrast, temperature variables contributed only between 1 - 1.87 %. The moderate contribution of agricultural land-use variables (e.g., sugarcane, cocoa, and cassava harvested areas) suggests that human-modified landscapes may influence transmission dynamics. We also investigated the relationship between the predictor variables and their impact on the prediction on OROV suitability. **Figure 1D** shows the distribution of SHAP values for each predictor variable, positive SHAP values indicate a contribution of the feature value in favour of positive OROV suitability and negative SHAP values are in favour of negative OROV suitability. For instance, we observe that humidity has a more positive impact on OROV suitability while water occurrence has more of a negative impact.

To better understand how environmental predictors influence OROV suitability, we visualized SHAP-based response curves alongside spatial response maps (See **Supplementary Figure S2**). The SHAP response curves depict how changes in each predictor influence OROV suitability, showing both the magnitude and direction of their effects, while the corresponding maps illustrate the spatial distribution of their influence across Brazil. Several variables exhibited strong non-linear effects. For example, humidity showed a sharp positive influence on OROV suitability beyond 70%, particularly across the Amazon Basin. Elevation demonstrated a negative association, with suitability decreasing rapidly above 250 meters, consistent with its lowland distribution. Soil moisture, banana harvest area, and forest loss also contributed positively in specific ranges, suggesting that moist, disturbed environments and agro-ecological transitions may favor virus circulation. In contrast, variables such as water occurrence and temperature extremes showed more complex or diminishing returns at higher values. The spatial patterns mirrored these effects. High SHAP values were clustered in the central and northern regions for variables like humidity, soil moisture, and cropland-natural vegetation, whereas temperature and elevation effects were more prominent in southern Brazil. These patterns highlight that environmental drivers of OROV suitability are both variable-specific and geographically structured, reinforcing the need to account for spatial heterogeneity when interpreting ecological suitability.

### Spatiotemporal Consensus Suitability Maps across Brazil

To obtain an overall OROV disease risk map for Brazil, we calculated a consensus map by integrating predictions from the best-performing models under each pseudo-absence sampling technique. We assigned ranks based on the BI score, converted them into weights by dividing each model’s rank by the sum of all ranks, and used these weights to generate a final weighted environmental suitability map, referred to as the consensus map. The consensus map of environmental suitability (**Figure 2A**) reveals spatial heterogeneity in OROV transmission potential across Brazil, with highly suitable regions concentrated in the Amazon Basin and along major river systems. The highest suitability is observed in northern and central regions, particularly in the states of Amazonas (AM), Pará (PA), Rondônia (RO), and Maranhão (MA). Notably, coastal regions of northeastern and southeastern Brazil also exhibit high suitability, suggesting that environmental conditions outside the traditional endemic zone may support virus circulation. Areas with low suitability are primarily located in the central-west and southern regions, where climatic and ecological conditions may be less conducive to sustained transmission.

**Figure 2.**
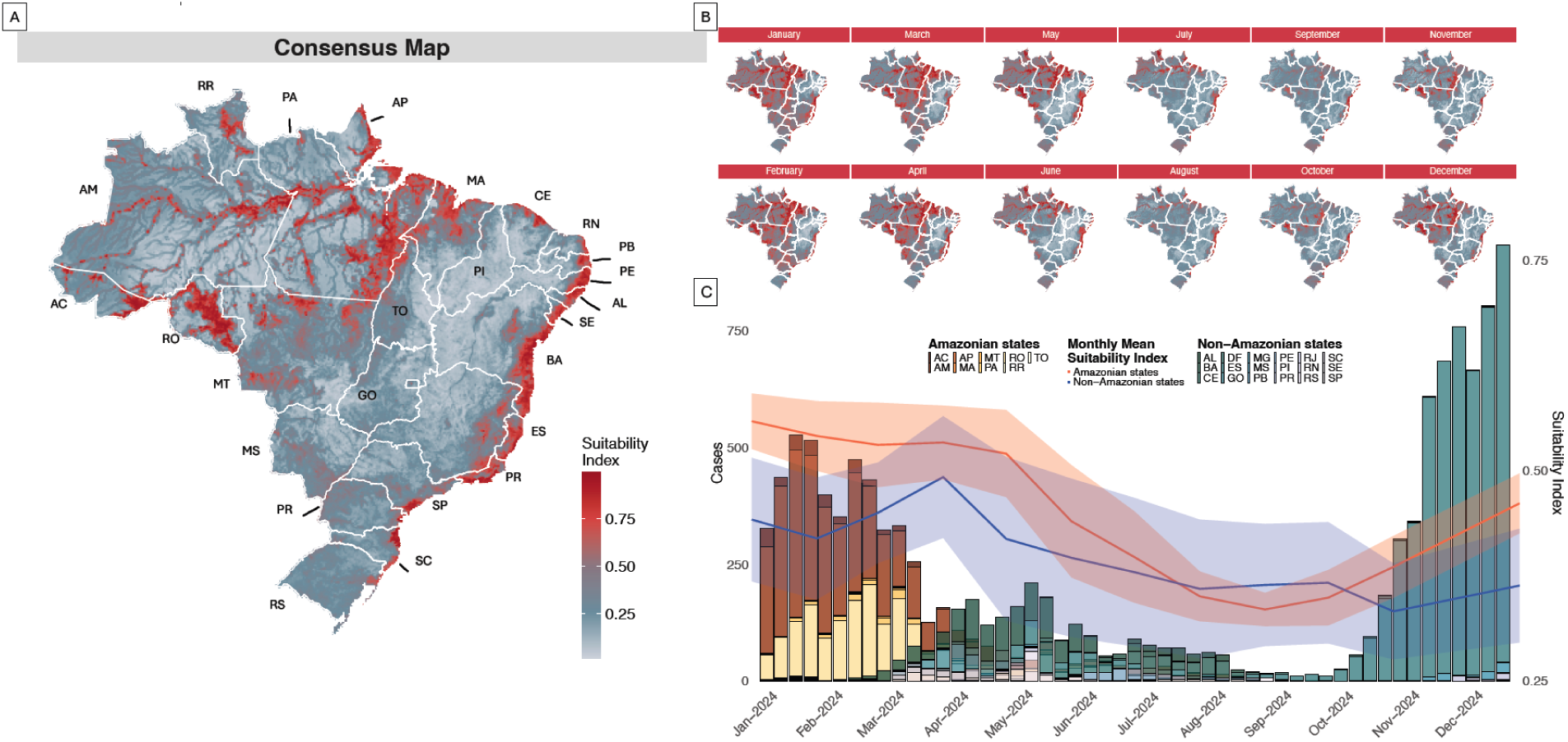
Consensus map and temporal environmental suitability trends. (A) The consensus map of environmental suitability for Oropouche virus transmission across Brazil, highlighting areas with persistent high suitability (red) and regions with lower suitability (blue). (B) Monthly projection of environmental suitability index across Brazil (maps) with temporal trends in case counts (stacked bars) and suitability index (lines) for the year 2024, with separate trends for Amazonian and non-Amazonian states. Shaded regions represent the variability in suitability estimates, with darker lines indicating mean suitability.

The suitability index was then projected over a monthly time interval in Brazil, considering variations in climatic variables. This temporal analysis (**Figure 2C**) highlights seasonal patterns in environmental suitability which closely align with reported case counts in the country. The suitability index is highest between January and May (>0.5) particularly in the Amazonian states, coinciding with the period of peak case numbers in Amazonian states. The decline in environmental suitability from June to September closely coincides with the drop in reported case numbers, suggesting that seasonal changes during this period create less favorable conditions for virus transmission. Towards the end of the year, environmental suitability begins to increase once again, coinciding with a rise in reported cases, particularly in non-Amazonian states. The monthly suitability maps (**Figure 2B**) further illustrate the dynamic seasonal shifts in transmission potential coinciding with increases and decrease of reported cases.

As a means of validating the seasonal pattern captured by our suitability model, we compared predicted monthly suitability to independently estimated reproduction numbers (Rt) derived from weekly OROV case data. We computed monthly Rt values separately for the Amazon and non-Amazon regions using a 4-week moving average (See **Supplementary Figure S3**), and correlated these with our monthly suitability predictions. The correlations were strong and statistically significant (Amazon: ρ = 0.75, p = 0.005; non-Amazon: ρ = 0.70, p = 0.018), indicating that the model effectively reflects seasonal variation in OROV transmission potential across distinct ecological settings.

### Continental Predictions of OROV and its primary vector, *C. paraensis,* across the Americas

We validated the model using independent OROV occurrence data from six countries across Central and South America (See **Supplementary Figure S4)**: Panama (n = 76), Ecuador (n = 3) ^24^, Colombia (n = 7)^25^, Cuba (n = 3) ^25^, Bolivia (n = 3) ^25^, and Peru (n = 21) ^24^, excluding Brazil, which was used for model training and internal validation. The model achieved a Boyce Index of 0.84, indicating strong predictive ability beyond the training region. The spatial predictions of OROV environmental suitability across the Americas reveal distinct high-risk regions concentrated in tropical and subtropical areas of South and Central America (**Figure 3A**). The model identifies high suitability along the Amazon Basin, the Atlantic Forest of Brazil, and portions of Colombia and Peru, reflecting known virus circulation areas. In Central America, suitability is notably high in Panama, Costa Rica, and parts of Honduras and Nicaragua, suggesting potential transmission risk beyond traditionally recognized endemic zones. Some coastal areas of Mexico and the Caribbean also exhibit moderate suitability, though these predictions are less pronounced **(Figure 3A)**. The United States of America exhibits moderate to low environmental suitability for OROV transmission, with only small, isolated areas of moderate suitability along the southeastern coastline and parts of the Gulf Coast.

**Figure 3.**
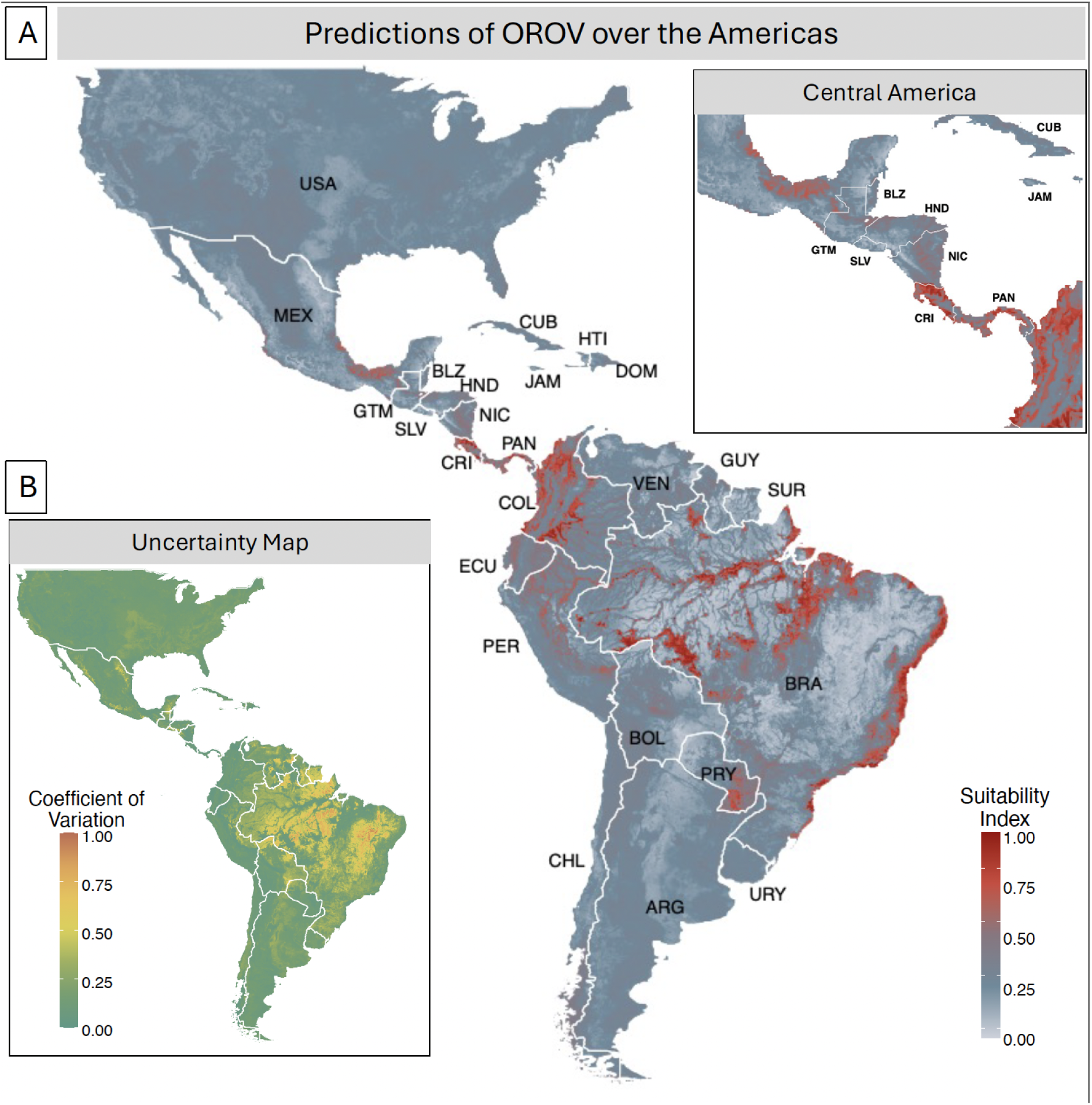
Predicted Environmental Suitability for Oropouche Virus (OROV), across the Americas. (A) Predicted environmental suitability for Oropouche virus transmission across the Americas. The inset highlights suitability predictions for Central America, including Panama (PAN), Costa Rica (CRI), Nicaragua (NIC), Honduras (HND), El Salvador (SLV), Guatemala (GTM), Belize (BLZ), Cuba (CUB) and Jamaica (JAM). (B) Uncertainty map displaying the coefficient of variation, across models built under different pseudo-absence sampling techniques.

The uncertainty map (**Figure 3B**) provides insight into the stability and consistency of model predictions across different pseudo-absence sampling strategies. The uncertainty values are derived as the variation across the five best-performing models used to build the consensus map. This variation quantifies the differences in predictions among these top models, highlighting areas where model agreement is high or low. Lower uncertainty (green areas) is observed in well-documented endemic regions, particularly in northern Brazil and part of the Amazon Basin, where high suitability predictions remain stable across all models. Conversely, higher uncertainty (orange to yellow regions) is present in parts of Central America, southern Brazil, and parts of the Andes, suggesting that predictions in these areas are more sensitive to the choice of pseudo-absence sampling technique.

We also built an ENM for the primary vector, *C. paraensis*, using the best-performing pseudo-absence sampling technique identified in the virus model evaluation. The Shapley analysis for the vector model (**Figure 4A**) highlights banana harvested area (8.21%) as the strongest predictor of the vector’s environmental suitability. Other important predictors include sugarcane harvested area (5.32%), humidity (3.41%),soil moisture (3.29%) and cassava harvested area (2.83%), reinforcing the role of agricultural landscapes and moisture availability in shaping the vectors’ environmental niche. Notably, temperature variables (1.75-2.11%) contributed marginally to the model. We also generated spatial response maps, to illustrate the positive or negative contribution of each predictor variable (See **Supplementary Figure S5**). We found that factors such as sugarcane harvest, cassava and banana harvest tend to affect the vector suitability positively in central and northeastern Brazil. In contrast, high elevation and low temperatures show negative SHAP values.

**Figure 4.**
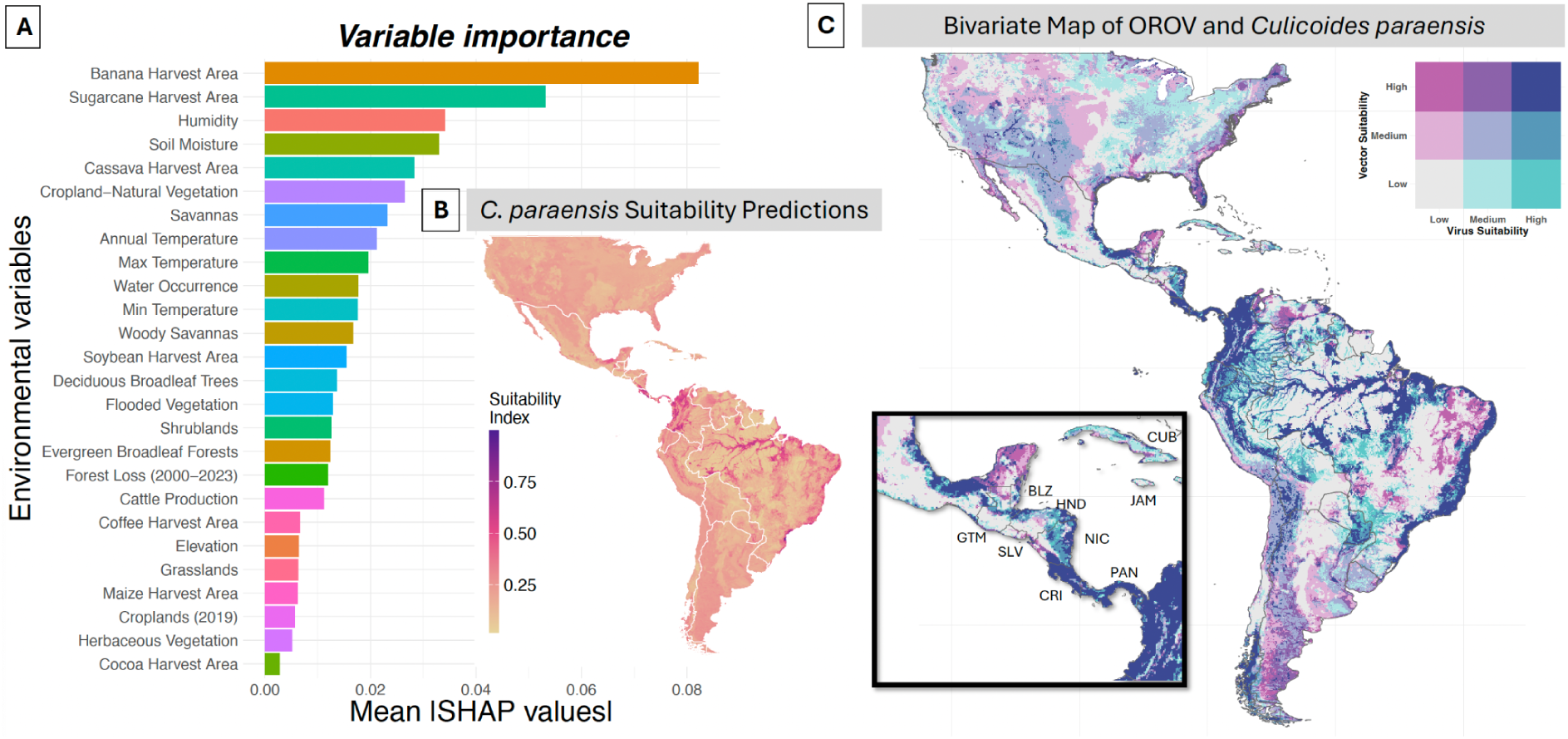
Environmental drivers and spatial suitability of *Culicoides paraensis* and bivariate suitability map of Oropouche virus (OROV) and its primary vector: (A) Variable importance plot showing the mean SHAP values for environmental predictors contributing to *C. paraensis* suitability. (B) Predicted spatial distribution of *C. paraensis* suitability across the Americas. (C) Bivariate suitability map of Oropouche virus (OROV) and*Culicoides paraensis* . The color gradient represents the combined suitability of OROV and each vector, with light gray indicating low suitability for both, pink/purple hues representing higher suitability for the vector, blue tones indicating higher suitability for the virus, and dark purple signifying high suitability for both. Inset highlights Central America and the Caribbean.

The vector’s suitability map (**Figure 4B**) mirrors a similar geographic pattern as the virus, particularly in Brazil, the Amazon Basin and parts of Central America. High suitability for the vector is observed in Central America, countries such as Costa Rica (CRI) and Panama (PAN) particularly in lowland and humid forested areas, aligning with the ecological preferences of *C. paraensis*. We also observe high environmental suitability areas concentrated in moist areas of Colombia and parts of the United states, particularly Florida. The uncertainty map for *C. paraensis* across the different pseudo absence runs exhibited higher divergence across the Amazon Basin (See **Supplementary Figure S6**).

### Exploring the overlap of viral-vector predictions

Given the similarities in spatial patterns, examining the bivariate suitability map of the viral and vector suitabilities together can provide insight into where both OROV and its vector are likely to co-exist and sustain transmission. We constructed a bivariate map depicting the relationship between OROV and its primary vector (**Figure 4C).** The bivariate maps reveal that regions with high suitability for both the virus and primary vector (*C. paraensis*) are concentrated in northern South America, Central America, and coastal Brazil, reinforcing the role of this vector in OROV transmission. Correlation analysis between the two maps indicates a strong association between the virus and its primary vector (r = 0.72). Although recent studies have detected OROV in Culex quinquefasciatus, no evidence to date supports its ability to transmit the virus. Emerging scenarios, such as the current epidemic in Cuba and molecular detection of the virus in Culex quinquefasciatus mosquitoes^4^, have led to speculation about a potential shift toward mosquito vectors; nonetheless, recent evidence does not support the competence of this species for OROV transmission^5^. Given these uncertainties, we mapped the spatial overlap between predicted OROV suitability and *Cx. quinquefasciatus* distribution (See Supplementary Figure S7), while emphasizing that such overlap does not confirm vector competence.

## 3. Discussion

Here, we present some of the first uses of ecological niche models application in OROV disease ecology and virus transmission. Unlike plants and animals, viruses depend on vector-host interactions, human movement and environmental persistence, making their spatial dynamics more complex^27,28^. Given these differences, we assessed whether traditional species modelling frameworks effectively capture virus distributions, focusing on the role of pseudo-absence selection and environmental drivers. Additionally, we investigated the environmental suitability of OROV alongside its primary vector, *C. paraensis*, and the potential role of *Cx. quinquefasciatus* as a secondary vector. In summary, we obtain a comprehensive continental assessment of the risks of transmission of this virus, which will be critical for public health planning.

In this study, for the OROV models, the target group-based sampling technique for pseudo-absences outperformed random sampling, particularly under small PA ratio^29^. The target group-based sampling technique produced the highest Boyce Index (BI = 0.96, ROC=0.904) with a small PA ratio and and the most spatially refined suitability predictions, supporting the idea that pseudo-absence selection strategies based on known virus occurrence records improve model accuracy^20^. Milanesi et al.^30^ reported that an observer-oriented approach using occurrences of a target group species as pseudo-absences improved ENM predictive accuracy compared to random pseudo-absences. Random sampling in our case resulted in relatively poor model accuracy across small PA ratios, which is a well-documented limitation in species distribution models when absences are not ecologically meaningful^31^. The strong performance of density-weighted sampling approaches suggests that incorporating demographic data into pseudo-absence selection improves the model’s ability to capture virus transmission patterns. While target group background sampling shows promise in addressing sampling bias and improving SDM performance, results may vary depending on the specific implementation and ecological context. Careful consideration should be given to defining appropriate target groups and the sampling domain, environmental heterogeneity, and resulting spatial predictions which is crucial for developing biologically realistic and fit-for-purpose habitat models ^30,32^.

The strong performance of both 1:1 PA ratios and 10,000 pseudo-absences suggests that model accuracy benefits from either balanced contrast or extensive background coverage. A balanced contrast (1:1 PA ratio) means that presence and pseudo-absence points are equally weighted, preventing biases that may arise when one class dominates the dataset. Extensive background coverage (10,000 pseudo-absences) allows the model to sample a wide range of environmental conditions, improving generalization by better representing areas where the virus is unlikely to occur^20,33^. In contrast, intermediate PA ratios (1:5, 1:10 with 2,000 – 4,000 pseudo-absences) do not provide enough contrast as 1:1 nor enough environmental representation as 10,000, leading to weaker model performance. While some PA ratios performed well under specific pseudo-absence sampling techniques, no single ratio consistently outperformed others across all approaches. This suggests that the most effective pseudo-absence ratio depends on the chosen sampling technique and the ecological or spatial characteristics of the target species or pathogen.

The SHAP analysis for variable importance (**Figure 1C & Figure 3C**) revealed notable similarities in the environmental drivers of OROV and *C. paraensis* suitability. Humidity emerged as the strongest predictor for OROV (7.08%), likely due to its role in vector survival, as it also influenced *C. paraensis* presence (3.41%). Land-use variables, particularly banana harvested area, were key predictors for both the virus and vector, possibly due to *C. paraensis*’ preference for breeding in decaying vegetation and agricultural waste^7,11,13^. Previous studies have reported that *C. paraensis* utilizes crop residues, such as banana and cocoa remains, as breeding sites in man-modified landscapes^7^. Our study found soil moisture to significantly influence the vector’s distribution (**Figure 3C**), which aligns with the vector’s lifecycle stages, including egg-laying, larval development, and adult survival^34^. As such, it serves as a hotspot for vector populations. Moreover the larvae of *C. paraensis* not only prefers moisture but also thrive in microhabitats such as tree holes, decaying organic matter whereby they are particularly drawn to rotting banana and plantain stalks^35^, which we also observed in this study (**Figure 3C**). While temperature is a well-established determinant of vector competence and viral replication in many mosquito-borne systems^36^, few studies have investigated the thermal ecology of *C. paraensis*. Given the limited number of presence records and potentially incomplete representation of the species’ full temperature niche in our dataset, the model may not capture the complete effect of temperature on vector suitability. Given the current gaps in vector occurrence data and the sparsity of ecological information for OROV, modeling viral presence directly allows us to capture broader environmental associations without relying solely on vector distributions, which may lead to over- or underestimation of risk in unsampled areas. More field and experimental studies are needed to better understand the ecology of *C. paraensis* and its role in OROV transmission.

These results highlight that high or low suitability for OROV is not driven by a uniform set of predictors across Brazil. Instead, different regions are influenced by distinct combinations of environmental factors. In the Amazon Basin, suitability is strongly shaped by humidity, elevation, and woody savannas. In contrast, in southern Brazil, variables like forest loss, sugarcane harvest, and temperature play a more prominent role. Our SHAP-based spatial response maps also show a strong influence of forest-related variables on OROV suitability (See **Supplementary Figure S2**). Forest loss contributes positively in much of the Amazon and northeastern Brazil, especially in actively deforested areas like the Arc of Deforestation. While the marginal response curve suggests that extreme forest loss may reduce suitability, the spatial SHAP patterns indicate that moderate disturbance is most often associated with increased risk. This supports the edge effect hypothesis, fragmented landscapes may enhance human-vector contact and provide suitable conditions for breeding and host presence. Evergreen broadleaf forest cover shows positive effects at intermediate levels, reinforcing the role of heterogeneous forest environments. In contrast, deciduous broadleaf cover has a more consistently negative effect, particularly in drier or open forest regions. Together, these patterns emphasize the ecological importance of land cover and disturbance in shaping OROV transmission risk.

The spatial and temporal dynamics of OROV suitability revealed in this study highlight the seasonal and geographic variability in transmission risk across Brazil. The consensus map (**Figure 2A**) indicates that high suitability is not limited to the Amazon Basin but extends along major river systems and into coastal regions of northeastern and southeastern Brazil, suggesting that environmental conditions outside traditionally recognized endemic zones may support virus circulation. This aligns with recent studies showing that vector-borne disease distributions are influenced by land-use change, deforestation, and urbanization, which can create new ecological niches for disease transmission^3^.

The seasonal fluctuation in suitability (**Figure 2B**) underscores the importance of climatic drivers in modulating OROV transmission potential. We found that the highest suitability and case numbers coincide with the rainy season (January–May 2024 in the Amazonian region) which aligns with previous studies that have observed Oropouche outbreaks occurring during the rainy season^2,8,13^, followed by a decline during the drier months (June–September), which likely results from reduced vector populations and lower environmental favorability for transmission (Feitoza et al., 2023). However, the resurgence of suitability towards the end of 2024, particularly in non-Amazonian states, suggests that seasonal transmission risk extends beyond traditionally endemic regions. This pattern may be driven by vector population dynamics, human movement, and changing land-use patterns, emphasizing the need for continuous surveillance beyond peak transmission months. The expansion of high suitability into southern and southeastern states by November–December 2024 highlights potential emerging hotspots, reinforcing the necessity of adaptive public health interventions that account for shifting transmission risks across both time and space.

The spatial predictions for OROV suitability indicate a strong alignment with known virus circulation areas, particularly across the Amazon Basin, Atlantic Forest, and portions of Central America (**Figure 3A**). However, the extension of high suitability into regions where OROV has not been extensively documented, such as southern Brazil and areas of the Andes in South America, suggests potential underreporting or conditions favorable for future virus expansion. Our virus suitability map also revealed a more widespread distribution in northern South America and parts of Central America exhibit high suitability for OROV. In Central America, coastal regions of Nicaragua, as well as parts of Costa Rica and Panama, exhibit high suitability for the virus. This observation also aligns with the suspicion that regions in Central America have a high number of unreported cases^2^. However, our principal component analysis (PCA) comparison between the modeling area and the prediction area (see **Supplementary Figure S8**) indicates that some environmental conditions in the prediction area extend beyond the range of the training data.

In contrast, *C. paraensis,* revealed a slightly more conservative distribution, with high suitability in Brazil, Columbia, Central America and coastal region of Mexico and Florida in the United States. This geographic distribution pattern of *C. paraensis* aligns with its preference for humid environments and its ability to thrive in urban areas^2,3^. The relationship between OROV and its vector is key to understanding transmission dynamics. While vector presence, environmental conditions, and human activity shape transmission risk (Ferraguti et al., 2021; Zouache et al., 2014), the geographic overlap between virus and vector suitability is not absolute. Areas with high OROV suitability but lower predicted vector presence suggest the potential involvement of a secondary vector, warranting further investigation into both the primary vector’s ecology and the possibility of alternative transmission routes.

Despite *C. paraensis* being recognized as the primary vector of OROV, the limited research on vector competence leaves gaps in our understanding of its full transmission cycle. A systematic review found that, in the six decades between OROV’s discovery and the 2023–24 outbreak, only seven vector competence studies had been published (Gallichotte et al., 2024). The recent OROV outbreak in Cuba, which coincided with a larger epidemic across the Americas, further raises questions about vector competence and transmission dynamics. Notably, Benitez et al. (2024) detected OROV RNA in *Cx. quinquefasciatus* mosquitoes, providing potential evidence of a secondary vector contributing to transmission. Our bivariate map also revealed that Cuba was at higher risk of OROV transmission with *Cx. quinquefasciatus* as a vector than *C. paraensis* (**Figure 4**). Given that virus distribution is shaped by both ecological and anthropogenic factors (Brunker et al., 2018; Finlay & Luck, 2011), further research is needed to assess the role of alternative vectors and environmental conditions that may facilitate virus persistence beyond its primary transmission pathways. Understanding how multiple vectors interact with OROV across different ecological settings is essential for refining targeted surveillance and control strategies (Ferraguti et al., 2021; Zouache et al., 2014), particularly in regions where virus and vector suitability do not fully align.

A potential limitation of this study is that our PCA comparing the modeling and prediction areas (refer to **Supplementary Figure S8**) reveals that certain environmental conditions in the prediction area fall outside the scope of the training data. This emphasizes zones with unique environmental characteristics and stresses the need for careful interpretation in these areas, as extrapolation may lead to increased uncertainty. Moreover, we make no differentiation between OROV lineages. In particular, the circulation of novel reassortant strains across South America could indicate differential adaptation to ecological niches based on viral genetic diversity^11,37^. Variants circulating in sylvatic versus urban environments may exhibit distinct environmental tolerances and transmission dynamics. This heterogeneity could significantly influence the virus’s spatial distribution. Additionally, little is known about the ecology of the primary vector, *C. paraensis*, with few studies investigating its geographic range, habitat preferences, or physiological constraints. Mechanistic laboratory experiments assessing vector competence, survival limits, and environmental tolerances remain scarce, limiting our ability to refine vector distribution models and fully understand its role in OROV transmission. Finally, a constraint common to many ecological niche modeling studies ^20,31,38^ is the lack of confirmed absence data for OROV, which constrains our ability to fully validate model predictions.

In conclusion, this study underscores the intricate relationship between environmental and climatic factors in shaping the distribution of OROV and its primary vector, *C. paraensis*. While evaluating different pseudo-absence sampling techniques and identifying key environmental predictors, we projected the spatial distribution of both the virus and vector across the Americas, identifying regions with high transmission potential and potential range expansions driven by climate and land-use changes. Additionally, we conducted monthly environmental suitability projections across Brazil, capturing seasonal fluctuations in OROV transmission risk, and we raise concerns about the possibility of transmission via secondary vectors. These findings provide valuable insights for refining predictive models and improving targeted surveillance and control strategies. A comprehensive understanding of these dynamics is essential for anticipating future OROV outbreaks and mitigating their impact on public health.

## 4. Methodology

### 4.1 Ecological Niche Modelling of Oropouche Virus

Ecological niche modelling (ENM) is a powerful approach for predicting the potential distribution of species and pathogens by identifying relationships between occurrence data and environmental variables^39^. Model selection generally depends on the type and quality of data available^40^. In this study, we apply ENM to assess the distribution of OROV using random forests (RF), a widely used machine learning technique known for its robustness in handling complex, high-dimensional ecological data^41^. RFs build a large ensemble of decision trees using random subsets of the training data, where each decision split is determined by a randomly selected subset of predictor variables^41–44^. The process starts by creating multiple bootstrapped samples, each containing the same number of data points, selected with replacement. Each sample is then used to train a classification tree, with splits based on the best predictor among a randomly chosen subset of candidate variables. Once the trees are fully trained (with a maximum of 500 trees), they are used to predict the ‘out-of-bag’ (OOB) samples - data points that were not included in the bootstrapped sample. These OOB samples are crucial for estimating the model’s accuracy and errors, and the misclassification rate of each tree is computed using OOB samples^42,45^.

ENMs typically rely on “background” or “pseudo-absence” data when true absence data is unavailable, using these as a contrast to presence locations in the model. A critical challenge in building the appropriate ENM is the selection of appropriate pseudo-absence data. To address this, we evaluate multiple pseudo-absence sampling strategies within the RF framework to determine the most suitable approach for modelling OROV’s environmental suitability distribution. The RF models are implemented using the R package biomod2 (version 4.2-5-2), allowing for a comprehensive assessment of how different pseudo-absence selection methods influence model performance. The code used for the analyses in this study can be found at [repository link].

#### 4.1.1 Virus Occurrence Data

Disease presence points of Oropouche were obtained and compiled from various locations in Brazil (molecular testing and sequencing records) from the years 1957 to 2024. Epidemiological data on OROV cases were retrieved from the Brazilian Ministry of Health, accessible at https://www.gov.br/saude/pt-br/assuntos/saude-de-a-a-z/o/oropouche. The data set includes confirmed case reports from all Brazilian states where OROV cases have been notified, including but not limited to Acre (AC), Alagoas (AL), Amazonas (AM), Bahia (BA), Ceará (CE), Minas Gerais (MG), Pará (PA), Rio de Janeiro (RJ), and São Paulo (SP). The data cover the years 2023 and 2024, organised by epidemiological week of reporting, and include key variables such as the municipality, state, and the year of occurrence. This data was geocoded at the municipality level and occurrence data were deduplicated by month. Additional OROV occurrence data was gathered and geocoded from all records in the GBIF (years: 1957-present) and Genbank (years: 2015-present) databases. After deduplicating all occurrence records, a total of 450 unique sampling location points remained (**Supplementary Figure S9A)**, covering the years 1957 to 2024, with the majority (∼85%) sampled in 2023-2024. For model development, the dataset was split into training and testing subsets, with 400 presence points used for training and randomly selected 50 presence points kept for model evaluation.

#### 4.1.2 Pseudo-absence sampling method

RF models require both presence and absence data to estimate the relationships between environmental predictors and species occurrence. However, true absence data are rarely available in ecological studies, particularly for infectious diseases and their vectors, due to the challenge of confirming species or pathogen absence at any given location. In the absence of true absence data, pseudo-absence points are generated and used in modelling. In this study, we compared the performance of five absence generation approaches:

**1. Random Sampling:** This method involves generating pseudo-absence points by randomly selecting locations within the study extent that did not overlap with presence points. This method assumes a uniform probability of species absence across the landscape, making it straightforward to implement.
**1. Geographic Sampling:** In this approach, pseudo-absences were generated within a defined geographic buffer or step-length around presence points. Unlike Random Sampling, this method first sets a spatial constraint around presence locations before randomly selecting pseudo-absence points within that area. By confining pseudo-absence points to areas near presence locations, the method accounts for the fact that species may be influenced by local environmental and spatial conditions, providing a more ecologically meaningful sampling strategy (See illustration in **Supplementary Figure S9-PanelB**) .
**1. Density-Weighted Geographic Sampling:** This method incorporates the density of occurrence points to guide pseudo-absence selection^46,47^. This method was tested both with and without the use of geographic buffers around presence points. By using the density of presence points as a proxy for sampling effort, pseudo-absences were more likely to be generated in areas with a higher concentration of presence observations. This reflects the reality that sampling is often biased toward accessible or frequently visited locations, the aim is to ensure that pseudo-absence points align with regions where sampling is likely to have occurred but the virus was not detected.
**1. Density-Weighted Population-Based Sampling:** Building upon the density-weighted geographic sampling approach, this method further refines pseudo-absence selection by integrating both the density of occurrence points and human population density. It was tested both with and without the use of geographic buffers around presence points, allowing for flexibility in accounting for spatial constraints. By incorporating population density, this approach reflects the critical relationship between human activity and the virus’ occurrence, particularly for species like OROV that depend on human hosts for transmission. The aim of this approach is to emphasise areas with higher human interaction where the species is absent but sampling efforts are likely.
**1. Target Group-Based Sampling:** Similarly, this approach, derived from Phillips et al.^48^, uses all occurrences of a predefined species group (the target group), in this case, other arboviruses such as dengue, chikungunya, yellow fever, and Zika^49^. The method uses the sampling density of these other arboviruses to guide the selection of pseudo-absence points, with the assumption that the sampling effort for the species of interest (OROV in this case) would follow a similar strategy. Importantly, the target group was chosen not based on ecological similarity or shared transmission pathways, but rather to approximate regions of historical arboviral surveillance activity. While transmission cycles differ, Aedes-associated arboviruses were prioritized due to their widespread testing in the region, serving as a practical proxy for the presence of arbovirus surveillance infrastructure. This approach was applied both with and without geographic buffers around presence points.

We evaluated those five pseudo-absence sampling strategies across five pseudo-absence ratios (PA ratio), four geographic buffer radii, and three Kernel Density Estimation (KDE) smoothing factors, leading to a systematic assessment of pseudo-absence selection effects

For each sampling strategy, we sampled pseudo-absences in the ratio of 1:1, 1:2, 1:5, 1:10 with respect to the presence points and a fixed number of 10,000 presence points across Brazil. For clarity, we will hereafter refer to this as the PA ratio. To account for spatial constraints in pseudo-absence selection, we investigated geographic buffers of 50–150 km, 50–300 km, and 50–500 km around presence points. A minimum buffer of 50 km was applied to ensure that pseudo-absence points were not placed in the exact same pixel as presence points.

For density weighted approaches, to quantify the spatial distribution of presence points, we applied Kernel Density Estimation (KDE) with a Gaussian kernel to generate a continuous density surface. The degree of smoothing is controlled by the bandwidth parameter (σ), which defines the spatial scale over which density is estimated. Given our Cartesian coordinate system, σ is expressed in degrees, where 1 degree ≈ 111 km. We evaluated three bandwidths (σ = 1, 3, and 5) to assess the impact of kernel smoothing. Regions with higher density indicate areas of greater sampling effort, guiding pseudo-absence selection to align with areas that were likely sampled but where the virus was not detected (See illustration in **Supplementary Figure S9C**).

The combinations of sampling strategies, PA ratios, buffer radii, and KDE smoothing factors resulted in 200 models (see **Supplementary Figure S10**). Each model was replicated three times with replacement to assess variability in pseudo-absence selection.

#### 4.1.3 Environmental Variables

In this study, we considered a total of 29 environmental variables, including climate-based and land-use variables. These environmental variables were chosen based on their known influence on vector habitat suitability and viral transmission dynamics^2,3,50^. After removing highly correlated variables (r > 0.8), except for maximum and minimum temperature to assess an optimum temperature effect, we retained 26 variables. All variables were in raster format, with all raster maps standardized to the lowest available resolution (∼27 km²) to ensure consistency across datasets. For climate variables, presence points were matched to their corresponding climate data based on the month of collection, ensuring temporal alignment between observed occurrences and environmental conditions at the time of sampling. The environmental variables were also acquired for the Americas (excluding Alaska and Canada) for projections outside of Brazil. Environmental variables for this study were sourced from publicly available remote sensing datasets and detailed information can be found in **Supplementary Table S1**.

### 4.2 Model Evaluation

To be able to compare the predictive performance of the ENMs built under different sampling techniques, we used the BI^51,52^. Given the lack of confirmed absence data for OROV, a common limitation in ecological niche modeling studies, we evaluated pseudo-absence sampling strategies by holding the presence data constant and varying only the absence selection. This allowed us to assess the discriminatory capacity of each method via the BI. The BI quantifies how well the predicted suitability values align with the actual occurrence points in the independent test dataset, making it particularly suitable for presence-only data. The BI is computed by comparing the frequency of predicted suitability values at observed presence locations (testing dataset) against the frequency of suitability values at randomly selected background points.

The index evaluates the relationship between habitat suitability predictions and observed presence points by comparing the predicted frequency of presence points (𝑃𝑖) to the expected frequency under random distribution (𝐸𝑖) across suitability bins (*i)*. The predicted-to-expected ratio (𝐹𝑖 = 𝑃𝑖 / 𝐸𝑖) is then computed for each bin. To reduce sensitivity to bin selection, the BI was computed using a moving window approach^52,53^. This method smooths the P/E curve by shifting a window across the suitability range, reducing biases from arbitrary binning. The Spearman correlation coefficient is calculated between these ratios and the midpoints of the bins, producing the BI. The index ranges from -1 to 1, with 0 indicating that the model is not different from a chance model. This estimate, derived from fixed bins, is referred to as the original Boyce Index (OBI).

Additionally, model performance was internally evaluated across different pseudo-absence replicates within each sampling strategy. The Random Forest models were evaluated using a block cross-validation approach to address spatial autocorrelation^54^. The study area was divided into four geographic blocks, designed to be of comparable size while also containing a minimum of 30 observations to maintain spatial independence between training and validation data. Models were trained on different combinations of these blocks and validated on the hold-out blocks. This method reduces the risk of overfitting and prevents performance inflation caused by spatial dependencies. We computed the True Skill Statistic (TSS) and the Receiver Operating Characteristic (ROC) curve for each model. These metrics were used to compare model performance and assess consistency across different sampling approaches. Full results, including TSS and ROC values, are provided in the Supplementary Materials (Annexe1).

#### 4.2.1 Variable Importance

To assess the contribution of each environmental variable to model predictions, we used SHapley Additive exPlanations (SHAP) values^55^, implemented via the itsdm R package^56^. SHAP values provide a consistent, model-agnostic framework for interpreting the influence of each predictor in complex machine learning models^55^. They estimate the marginal contribution of each predictor by considering all possible combinations of variables and attributing the change in prediction to each variable accordingly. A positive SHAP value indicates that the variable increases predicted suitability at a given location, while a negative value suggests it decreases suitability. This approach enables both the direction and magnitude of a variable’s influence to be quantified for each prediction. We computed SHAP values for each observation in the test dataset and then summarized the mean absolute SHAP value across all observations for each variable to estimate its overall importance. Variables with higher mean SHAP values were considered more influential in driving model predictions. In addition to summary importance values, we also generated spatial response maps to visualize how each variable influenced predictions across space. These maps display the SHAP value associated with each variable at each location, highlighting where a variable contributes positively or negatively to predicted suitability.

#### 4.2.2 Monthly Projections and Predictions across the Americas

After generating models from the different pseudo-absence sampling techniques, we evaluated their performance using the BI and ranked them accordingly. The best-performing model from each sampling technique was selected and assigned a rank based on its BI score. Previous studies have demonstrated that ensemble modeling, which integrates predictions from multiple statistical and machine learning algorithms, improves model performance by accounting for variability across different modeling approaches^57,58^. In this study, we extend this concept by constructing a consensus environmental suitability map from the different pseudo-absence sampling techniques rather than different modeling algorithms.

To construct a consensus prediction, we calculated a weight for each model according to its rank. The model with the highest BI score received the highest rank (5), while the lowest-performing model received the lowest rank (1). We then converted these ranks into weights by dividing each model’s rank value by the sum of all ranks, ensuring that higher-performing models contributed more to the final prediction. These weights were then used to generate a final weighted environmental suitability map, integrating information from all sampling techniques, which, for simplicity, we refer to as the consensus map.

For monthly projections in Brazil, we produced suitability maps based on monthly 2024 climate variables, while keeping all other environmental predictors at their yearly values. This allowed us to capture seasonal transmission patterns influenced by climatic fluctuations. For temporal comparison with OROV recorded cases in Brazil, we computed the monthly mean environmental suitability index separately for Amazonian and non-Amazonian states to assess how suitability trends align with observed case fluctuations. To evaluate the seasonal relevance of our environmental suitability predictions, we also computed monthly effective reproduction number (R_t_) values for OROV as an independent validation metric. R_t_ represents the average number of secondary infections generated by a single case over time and was calculated separately for Amazonian and non-Amazonian regions using weekly reported case counts, smoothed with a 4-week moving average. We then assessed the correlation between monthly R_t_ values and predicted environmental suitability across regions to evaluate whether temporal patterns in modeled suitability aligned with observed transmission dynamics.

For larger spatial-scale predictions, we applied the consensus model using mean annual climate variables to generate an overall environmental suitability map for the Americas (excluding Alaska and Canada). We validated the model using independent OROV geocoded occurrence data from six countries across Central and South America (see Supplementary Figure S4): Panama (n = 76), Ecuador (n = 3)^24^, Colombia (n = 7) ^25^(Fischer et al., 2025), Cuba (n = 3)^25^ (Fischer et al., 2025), Bolivia (n = 3)^25^, and Peru (n = 21)^25^, excluding Brazil, which was used for model training and internal validation. Geographic locations in Panama were recorded as part of an outbreak response in Panama at the beginning of 2025. We also performed a Principal Component Analysis (PCA) to compare the environmental niche of the modeling area with that of the prediction area. This allowed us to evaluate the extent of overlap and identify regions where predictions may extend beyond the range of sampled environmental conditions, providing insight into potential extrapolation uncertainty.

4.3 Environmental Suitability of *Culicoides paraensis*, the primary Vector of OROV

For the occurrence data of *C. paraensis*, the primary vector of OROV, we utilised geocoded records obtained from the Global Biodiversity Information Facility (GBIF, 2024) as well as data from several published studies^3,59–61^. After data cleaning, including duplicate removal and georeferencing verification, 78 occurrence points from Brazil were retained. However, this limited number of records may not fully capture the species’ ecological range, making comprehensive modeling challenging.

We fitted a RF model using the target group-based approach for pseudo sampling based on the best performing model for OROV. Presence data from all species within the Ceratopogonidae family, to which *C. paraensis* belongs, were used to construct a sampling density map, assuming that the distribution of *C. paraensis* overlaps spatially with other closely related species. A total of 1,862 occurrence points for the Ceratopogonidae family were retrieved from GBIF (1945–2024), and after data cleaning, 286 uniquely geocoded records were retained. These points were not included as presence points in modelling *C. paraensis* but solely to guide the selection of pseudo absence points by approximating where related species have been surveyed.

To explore the potential overlap in environmental suitability between OROV and its primary vector, we generated a bivariate suitability map to visualise the co-suitability of OROV and *C. paraensis*. We also explore the potential overlap with the suspected secondary vector, *Cx. quinquefasciatus*. A comprehensive suitability map for *Cx. quinquefasciatus* was obtained from Mendonça et al.^5^, allowing us to assess potential regions where both primary and secondary vectors may facilitate OROV transmission.

## Data Availability Statement

Current and future climate data are available at https://cds.climate.copernicus.eu/datasets. Anthropogenic data are available at https://www.earthdata.nasa.gov/. Data and code used in this study can be found at: https://github.com/CERI-KRISP/OROV_Vector_vs_Virus_Modelling

## Declaration of interests

We declare no competing interests.

## Contributors

J.P. and H.T. conceptualised and designed the study. J.P. analysed data, executed all primary data visualisations, and wrote the original draft. G.D. contributed towards the data extraction. M.G., L.A. and V.F. provided disease occurrence data. H.T., M.D., J.P., M.U.G.K, O.J.B. and A.L. contributed to methods development. C.B. and T.d.O. acquired funding for this study. H.T., M.D. and T.d.O supervised this work. All authors reviewed the manuscript.

## Supporting information

Supplementary Annexe 1

## Acknowledgements

CERI and KRISP are supported in part by grants from the Rockefeller Foundation (HTH 017), the National Institute of Health USA (U01 AI151698, also M.G.) for the United World Antiviral Research Network (UWARN), the Abbott Pandemic Defense Coalition (APDC), the SAMRC South African mRNA Vaccine Consortium (SAMVAC), the Medical Research Foundation (MRF-RG-ICCH-2022-100069), the Wellcome Trust (228186/Z/23/Z), and the Novo Nordisk Foundation (NNF24OC0094346, also M.G. and M.U.G.K.). O.J.B. was supported by a UK Medical Research Council Career Development Award (MR/V031112/1) which also supports AL. M.U.G.K. acknowledges funding from The Rockefeller Foundation (PC-2022-POP-005), Google.org, the Oxford Martin School Programmes in Pandemic Genomics & Digital Pandemic Preparedness, European Union’s Horizon Europe programme projects MOOD (#874850) and E4Warning (#101086640), Wellcome Trust grants 303666/Z/23/Z, 226052/Z/22/Z & 228186/Z/23/Z, the United Kingdom Research and Innovation (#APP8583), the Medical Research Foundation (MRF-RG-ICCH-2022-100069), UK International Development (301542-403), the Bill & Melinda Gates Foundation (INV-063472) and Novo Nordisk Foundation (NNF24OC0094346). The content and findings reported herein are the sole deduction, view and responsibility of the researcher/s and do not necessarily reflect the official position and sentiments of the funding agencies.

## Funding Statement

The funders had no role in data collection, analysis, interpretation of data, writing of the manuscript, or the decision to submit it for publication.

## 5. Supplementary Materials

**Figure S1.**
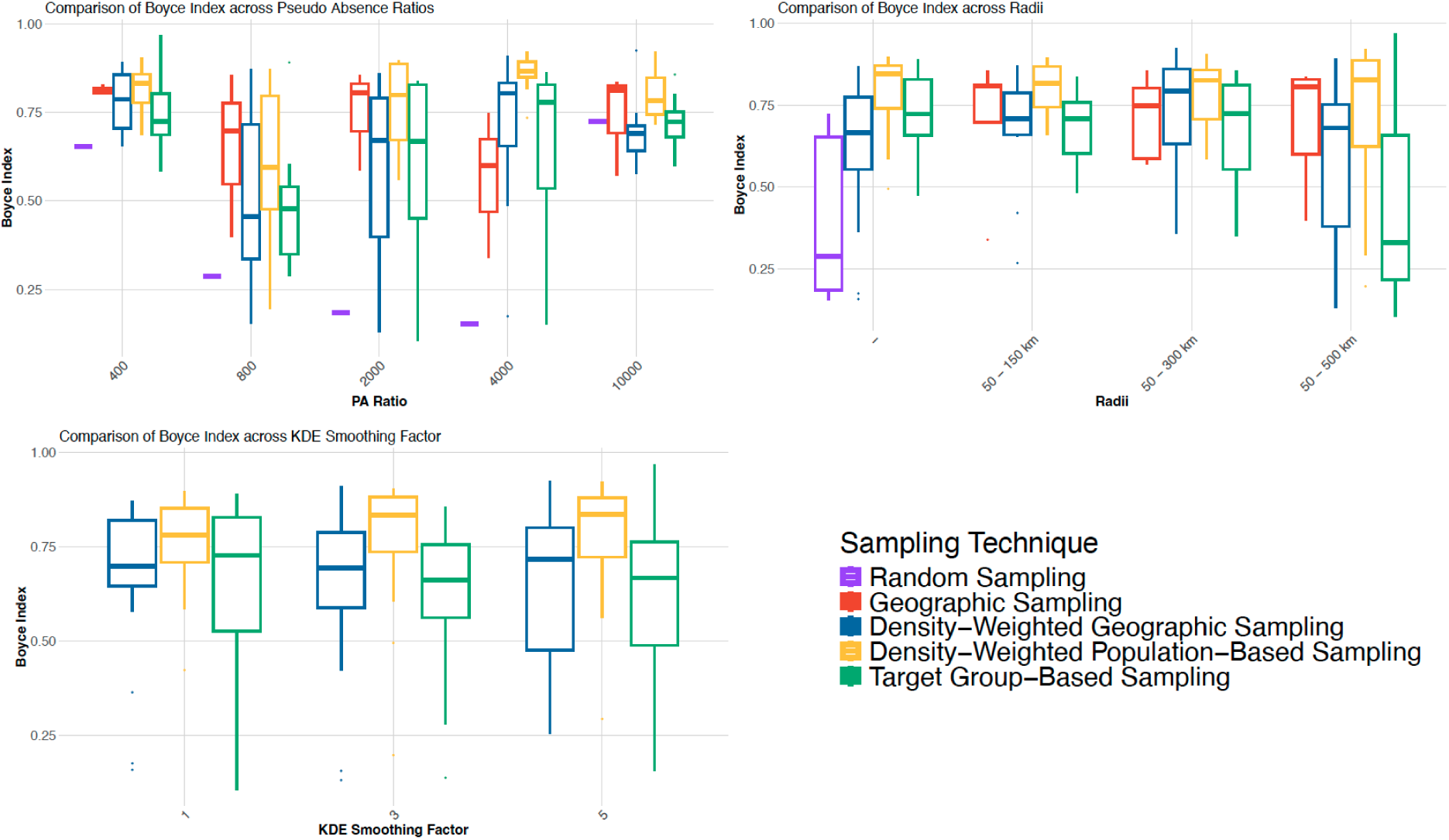
**Comparison of Boyce Index across different pseudo-absence sampling strategies**, evaluated using (A) pseudo-absence ratios, (B) geographic buffer radii, and (C) kernel density estimation (KDE) smoothing factors. The boxplots display the distribution of Boyce Index values for each sampling strategy, with higher values indicating better model performance. Sampling techniques include Random Sampling (purple), Geographic Sampling (red), Density-Weighted Geographic Sampling (blue), Density-Weighted Population-Based Sampling (yellow), and Target Group-Based Sampling (green).

**Figure S2:**
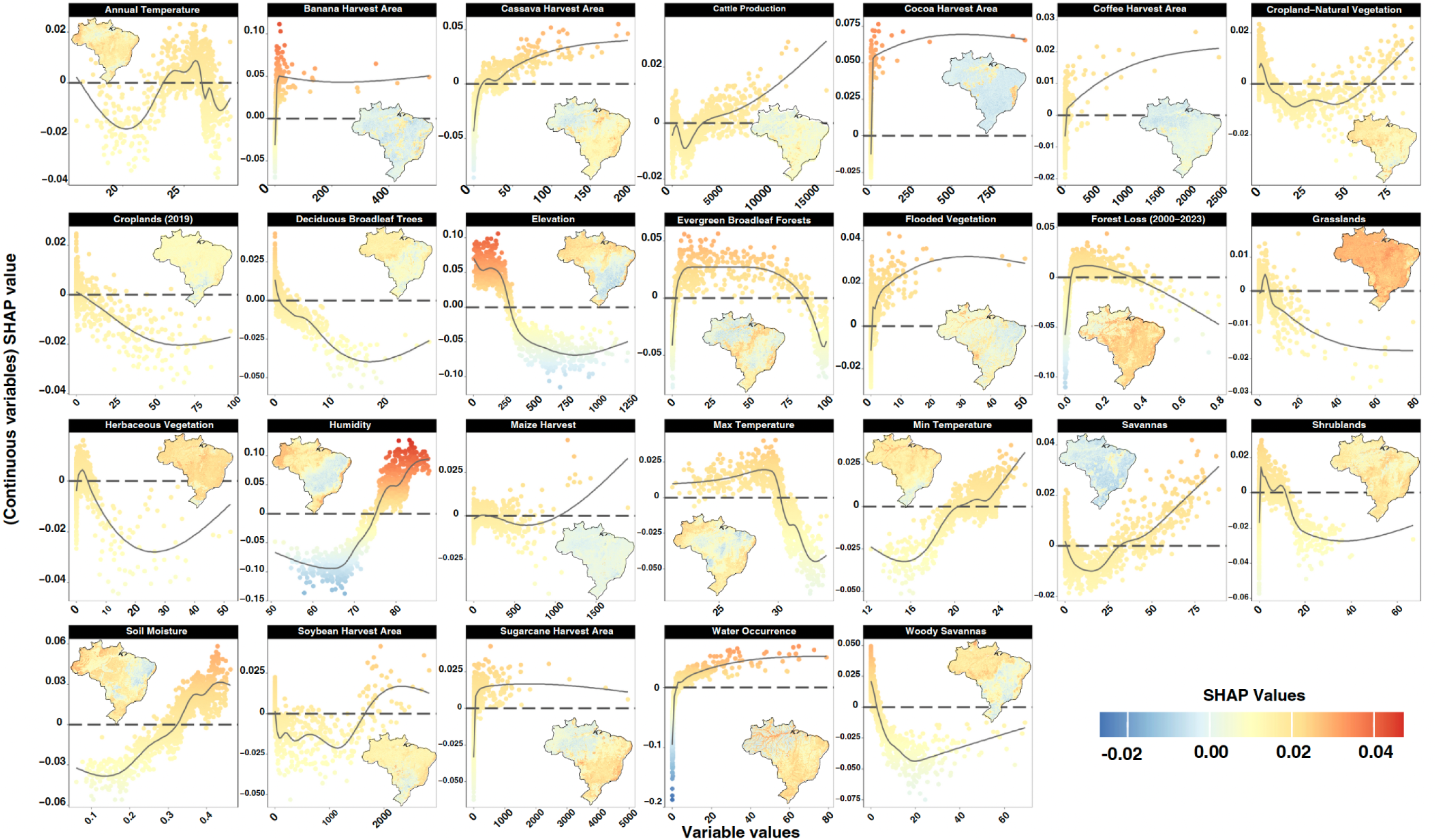
OROV Spatial Response Curves and Maps across Brazil for each environmental predictor variable. Response curves (scatter and smoothed lines) illustrate the direction and magnitude of each variable’s effect on model predictions. Maps show the spatial distribution of SHAP values, highlighting areas where each variable contributes positively (red) or negatively (blue) to predicted suitability.

**Figure S3:**
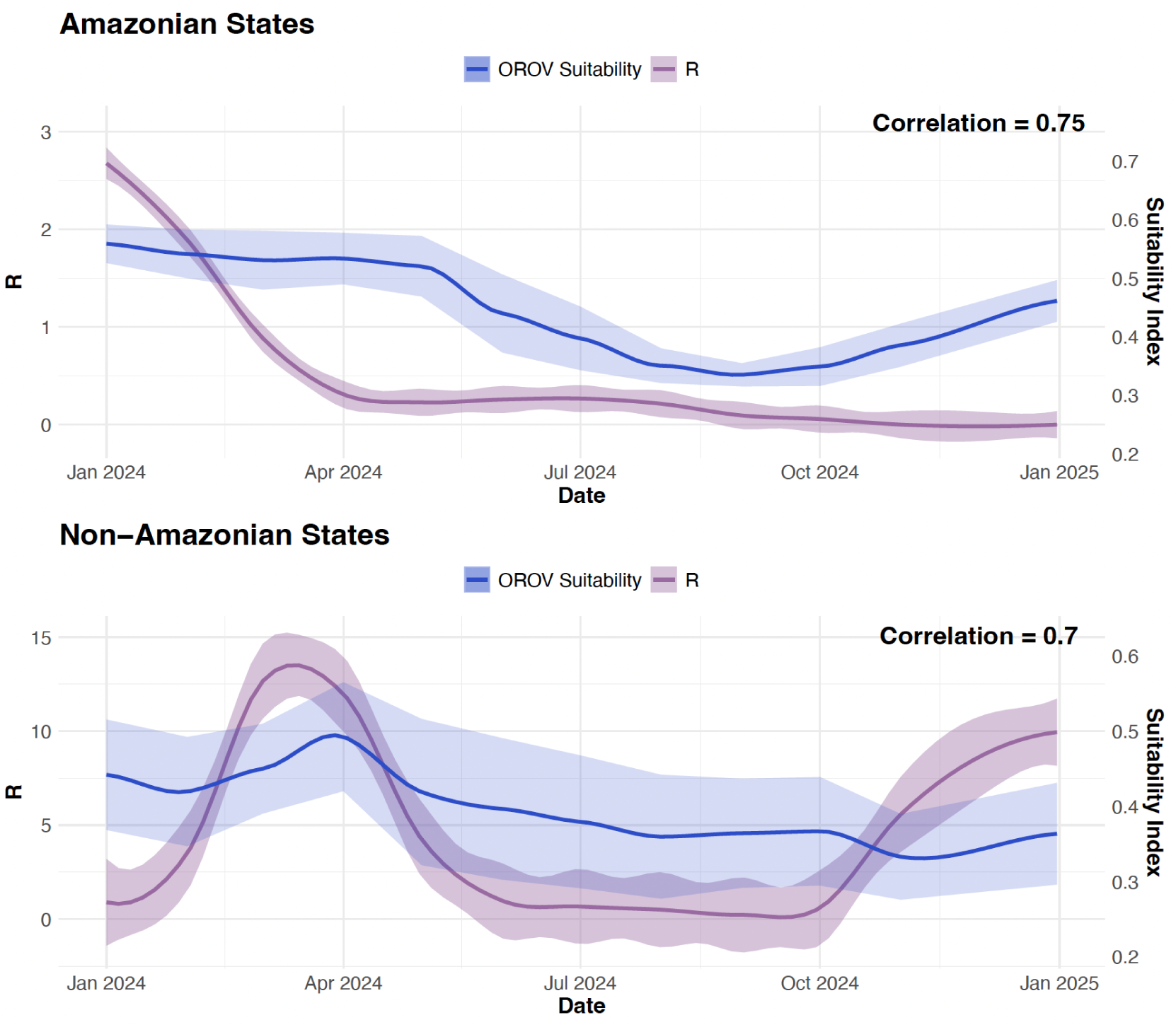
Estimated time-varying reproduction number (Rt) for Oropouche virus (OROV) in Brazil, stratified by region. (A) Amazon Region: includes the states of Acre, Amazonas, Amapá, Pará, Rondônia, Roraima, Tocantins, and Maranhão. (B) Non-Amazon Region: includes all other Brazilian states with reported OROV cases outside the Amazon basin. Shaded areas represent 95% confidence intervals.

**Figure S4:**
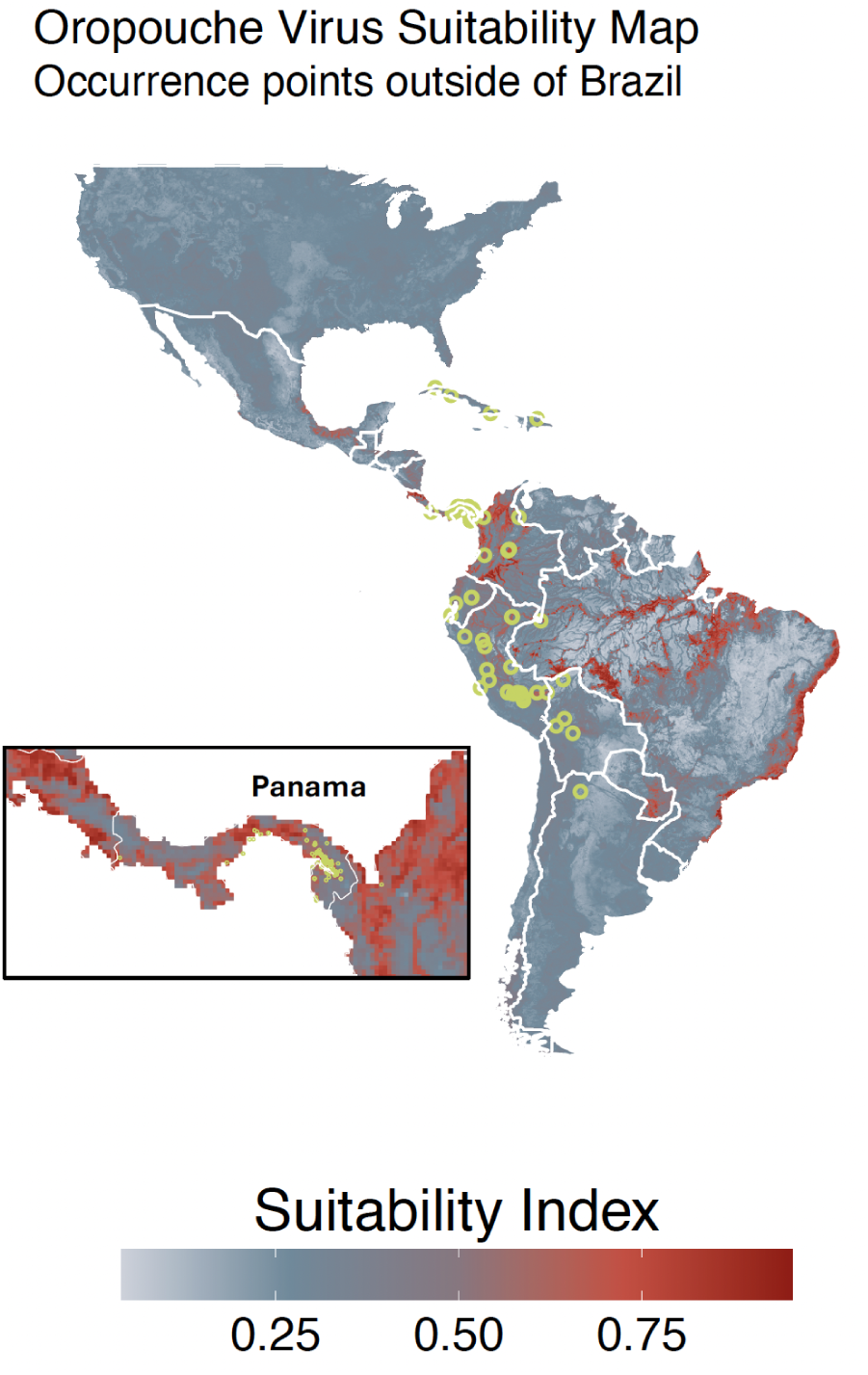
Predicted OROV environmental suitability across Central and South America, excluding Brazil. Green points represent independent geocoded occurrence locations used for model validation (n = 113), with an inset highlighting high-resolution results for Panama (n = 76).

**Figure S5:**
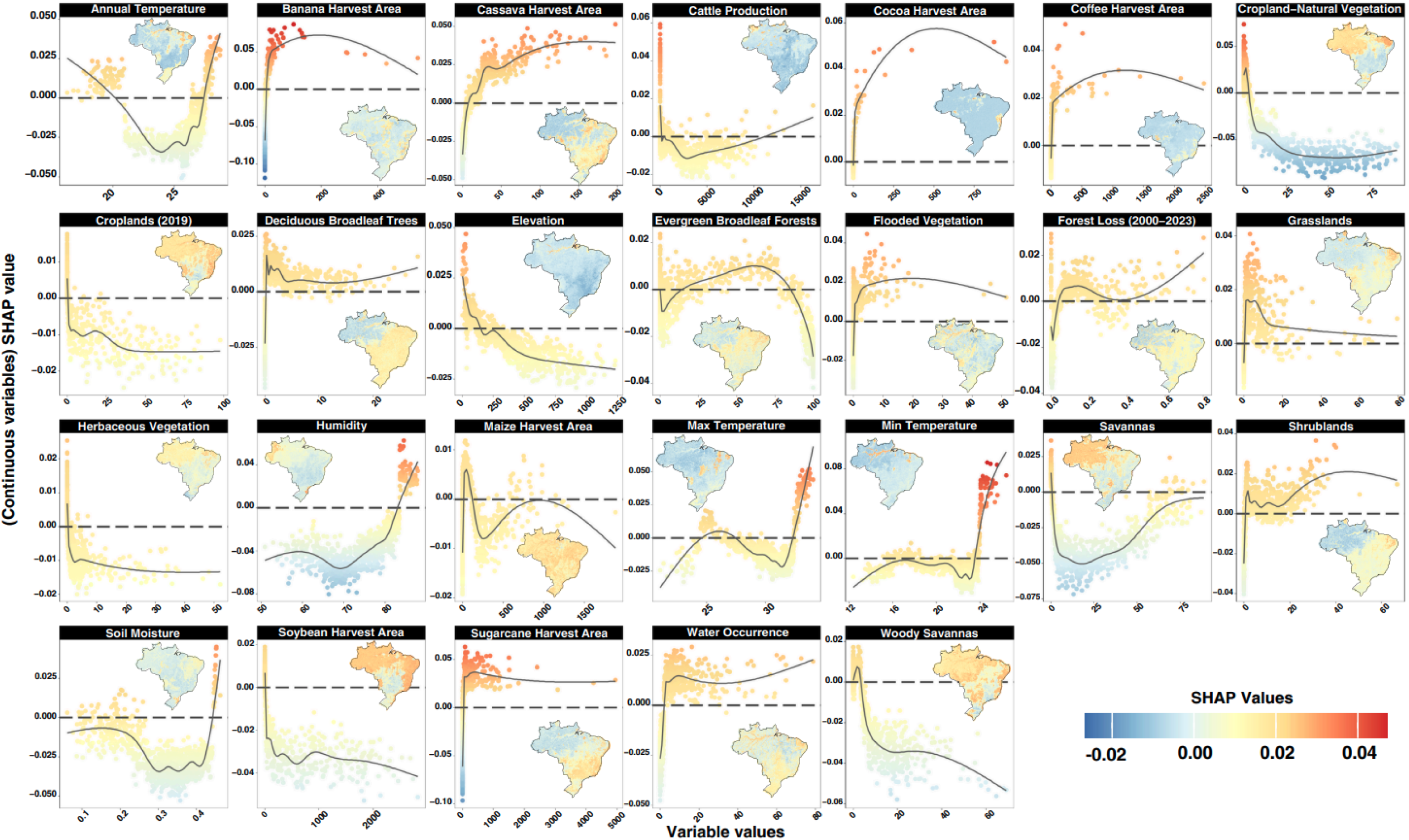
***Culicoides paraensis* Spatial Response Curves and Maps across Brazil for each environmental predictor variable.** Response curves (scatter and smoothed lines) illustrate the direction and magnitude of each variable’s effect on model predictions. Maps show the spatial distribution of SHAP values, highlighting areas where each variable contributes positively (red) or negatively (blue) to predicted suitability.

**Figure S6:**
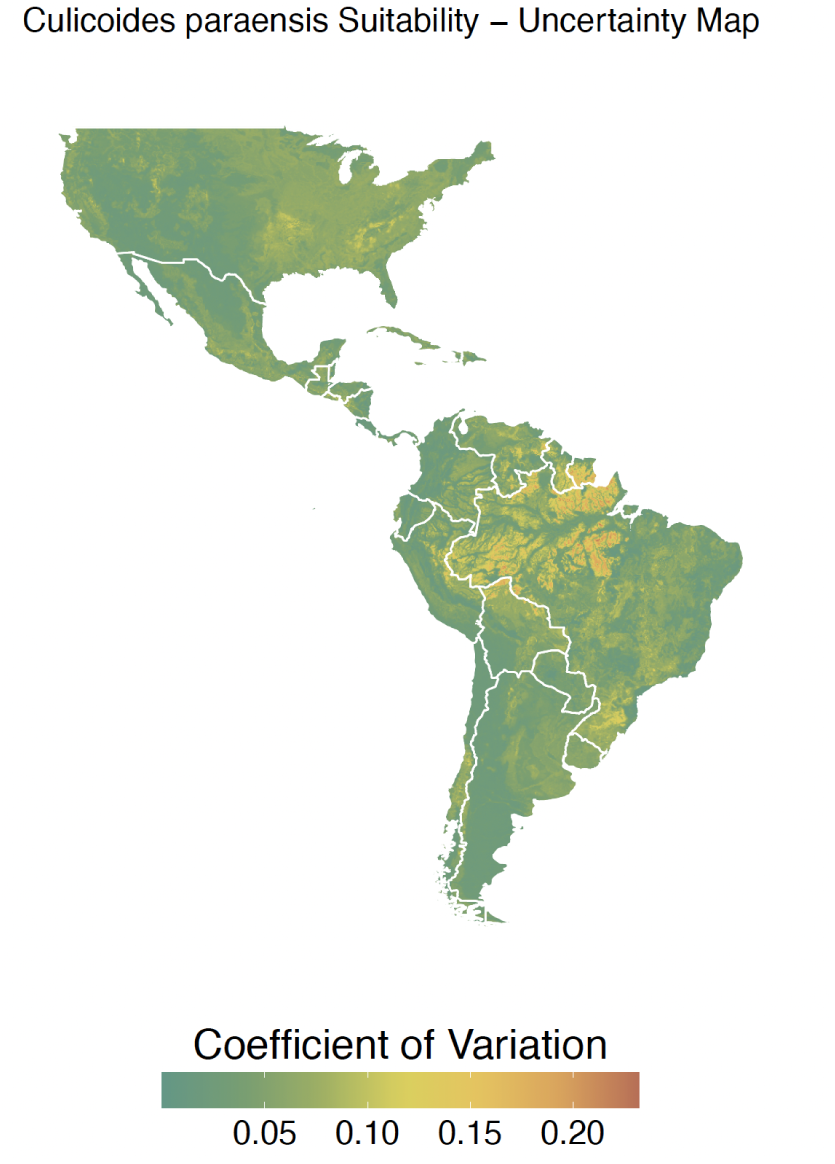
Uncertainty map for *Culicoides paraensis* suitability distribution, displaying the variation, across different pseudo absence runs for the target-based approach.

**Figure S7:**
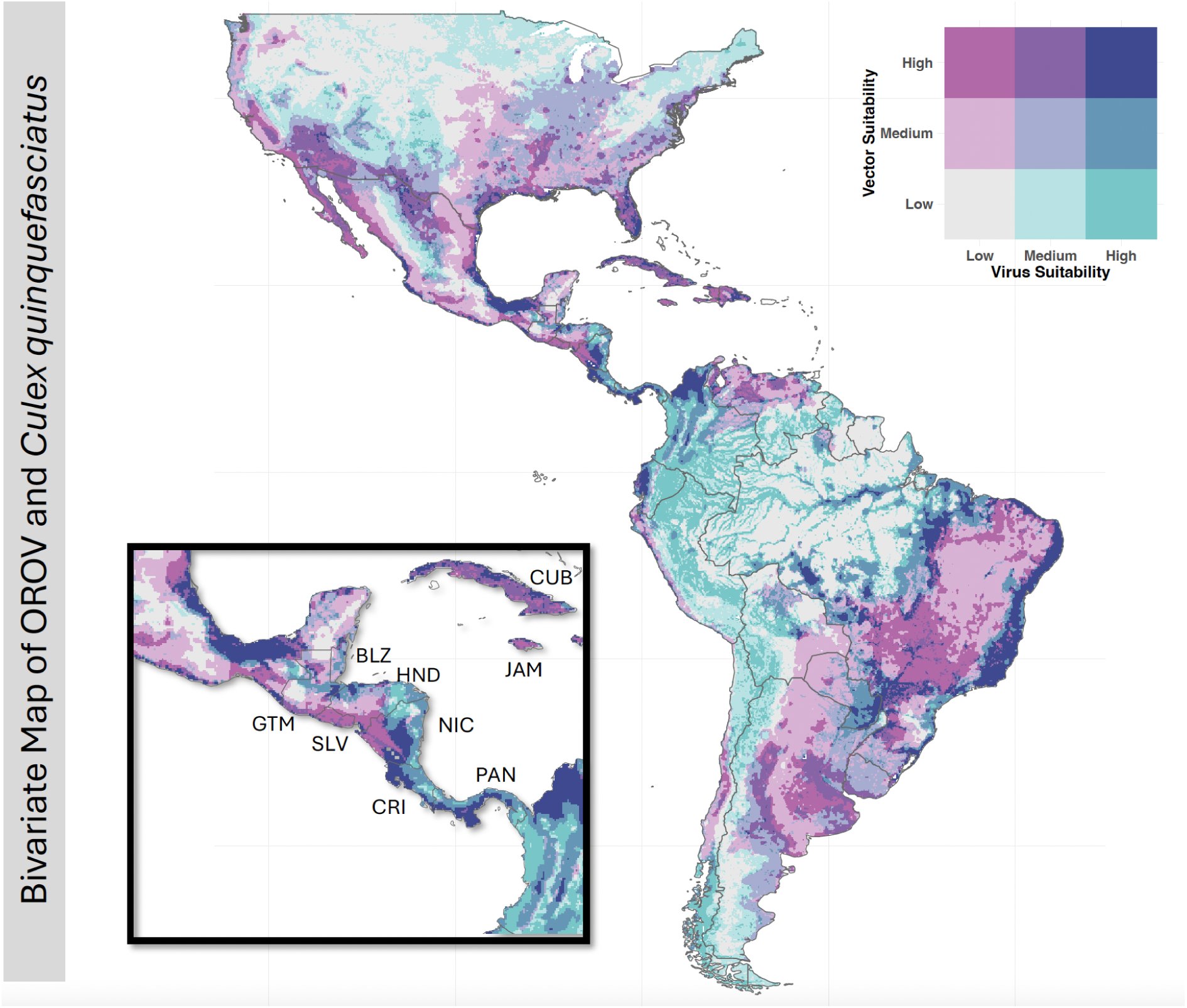
Bivariate suitability map of OROV and suspected secondary vector *Culex quinquefasciatus* (data obtained from ^5^)

**Figure S8.**
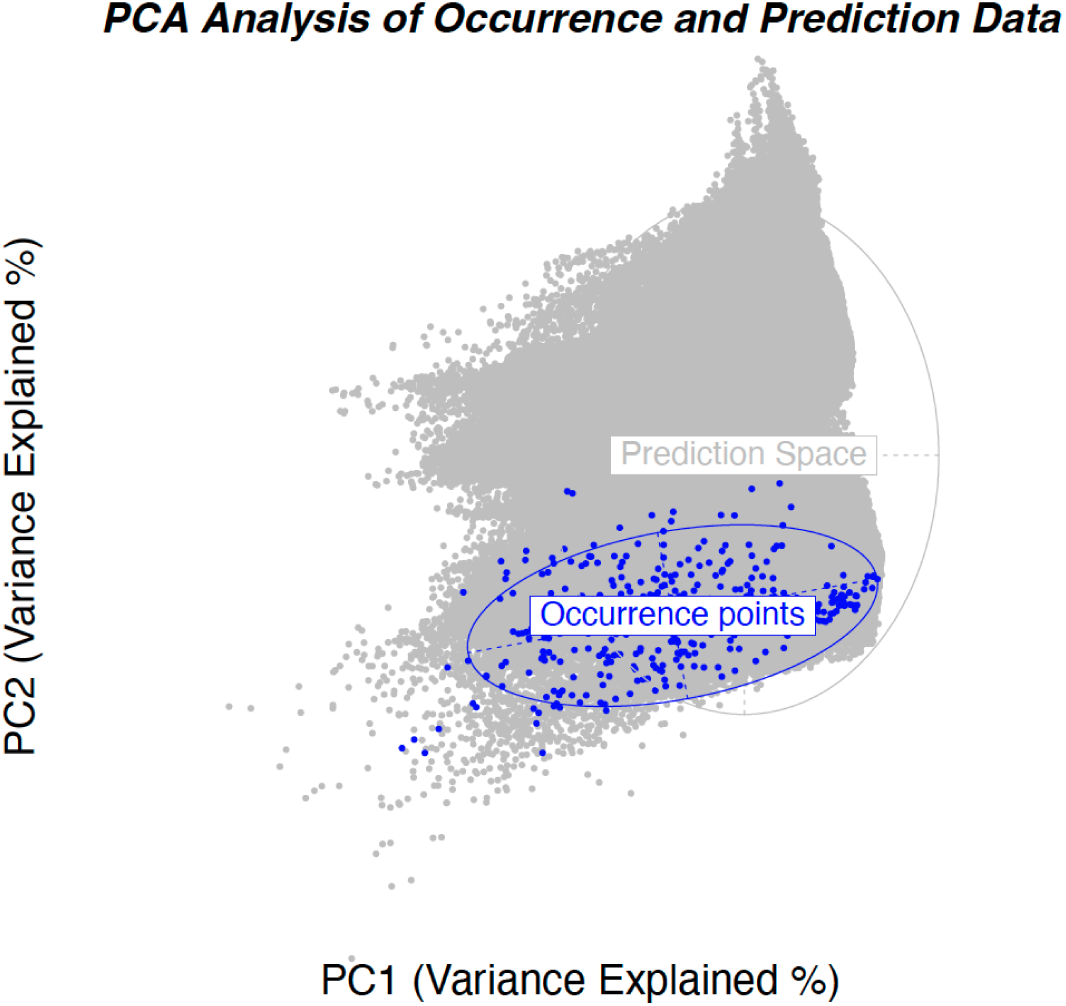
Principal Component Analysis (PCA) comparing environmental conditions in the occurrence points (blue) and the prediction space (grey). The PCA was conducted using the 26 environmental variables included in the model. The blue ellipse highlights the environmental conditions associated with observed occurrences, while the grey background represents the broader environmental space where predictions were made.

**Figure S9.**
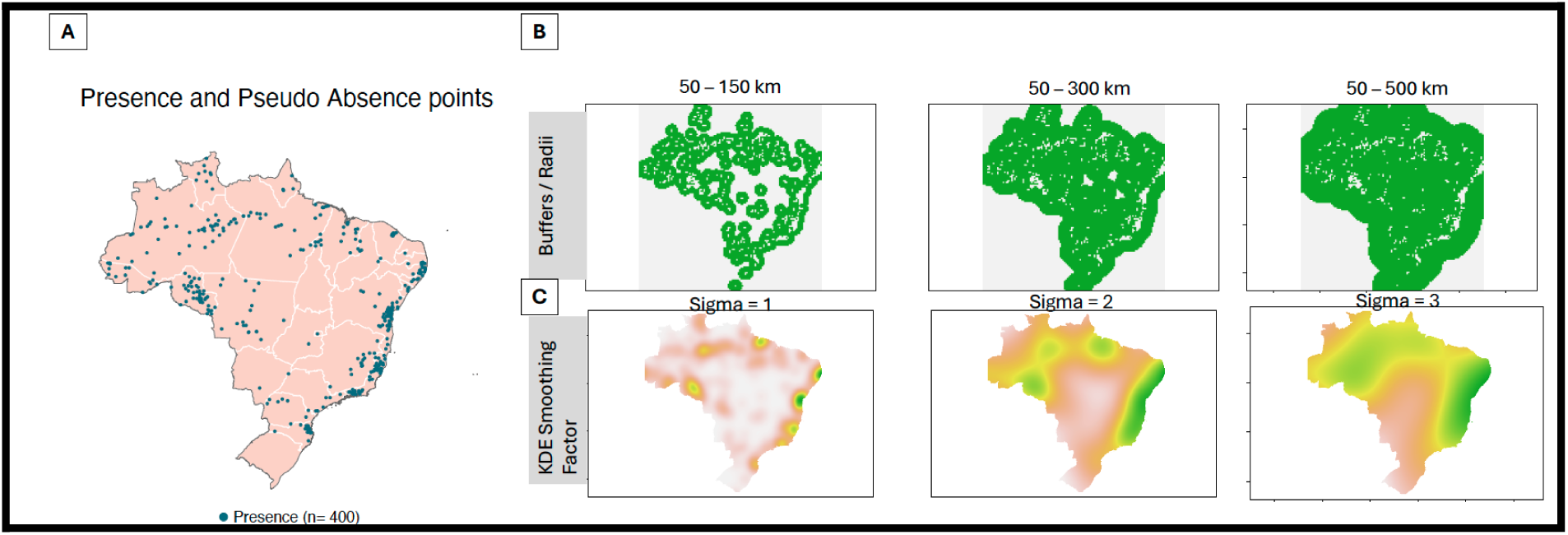
Illustration of spatial distribution of presence and pseudo-absence points used for modeling. (A) Map of 400 presence points (blue) distributed across Brazil. (B) Pseudo-absence selection using geographic buffers around presence points, with three buffer radii: 50–150 km, 50–300 km, and 50–500 km. Increasing the buffer size results in a broader spatial distribution of pseudo-absence points (green). (C) Kernel Density Estimation (KDE) smoothing factors (σ = 1, 2, 3) used to weight pseudo-absence selection based on occurrence point density. Higher σ values result in more spatially generalized pseudo-absence distributions. [Please note this is for illustration purposes].

**Figure S10.**
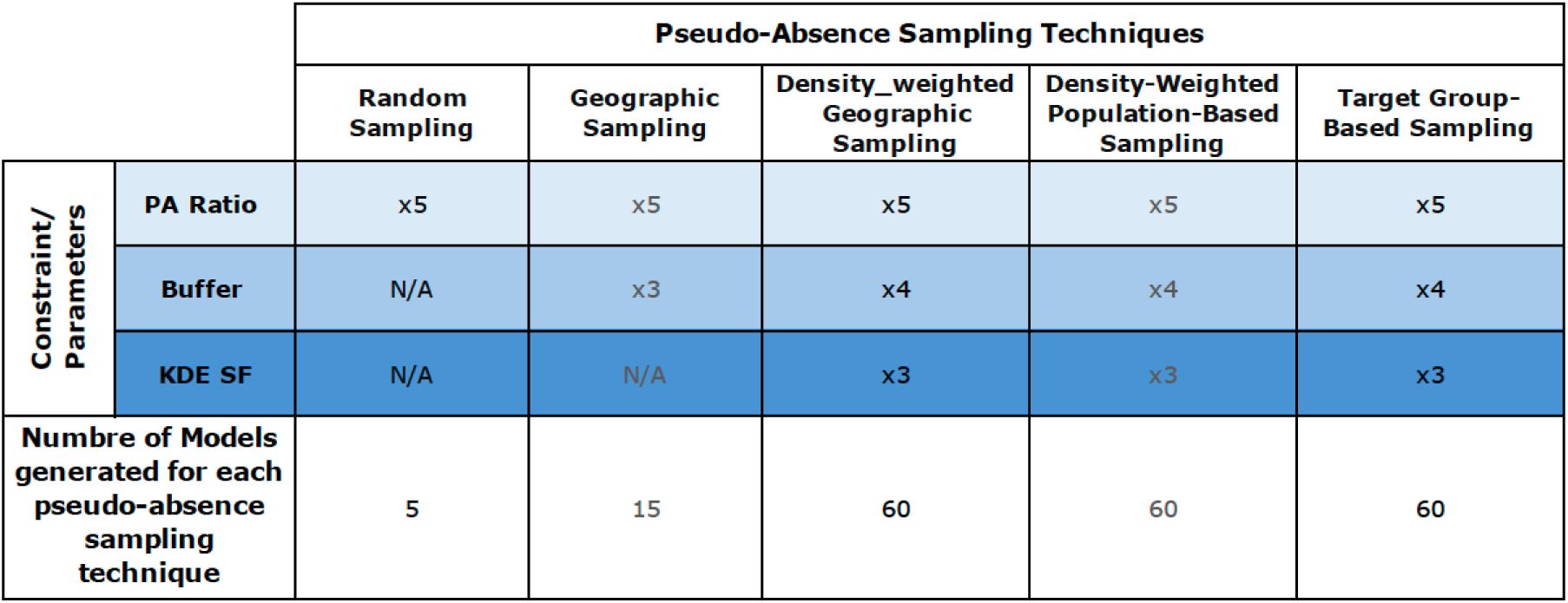
**Summary of the number of models generated under each pseudo-absence sampling technique**. Models were generated based on combinations of pseudo-absence (PA) ratios, geographic buffer radii, and Kernel Density Estimation (KDE) smoothing factors (σ). The table shows how different factors were applied across five sampling techniques: Random Sampling, Geographic Sampling, Density-Weighted Geographic Sampling, Density-Weighted Population-Based Sampling, and Target Group-Based Sampling. The final number of models reflects the total combinations of PA ratio, buffer size, and KDE smoothing factor used for each method.

**Table S1:**
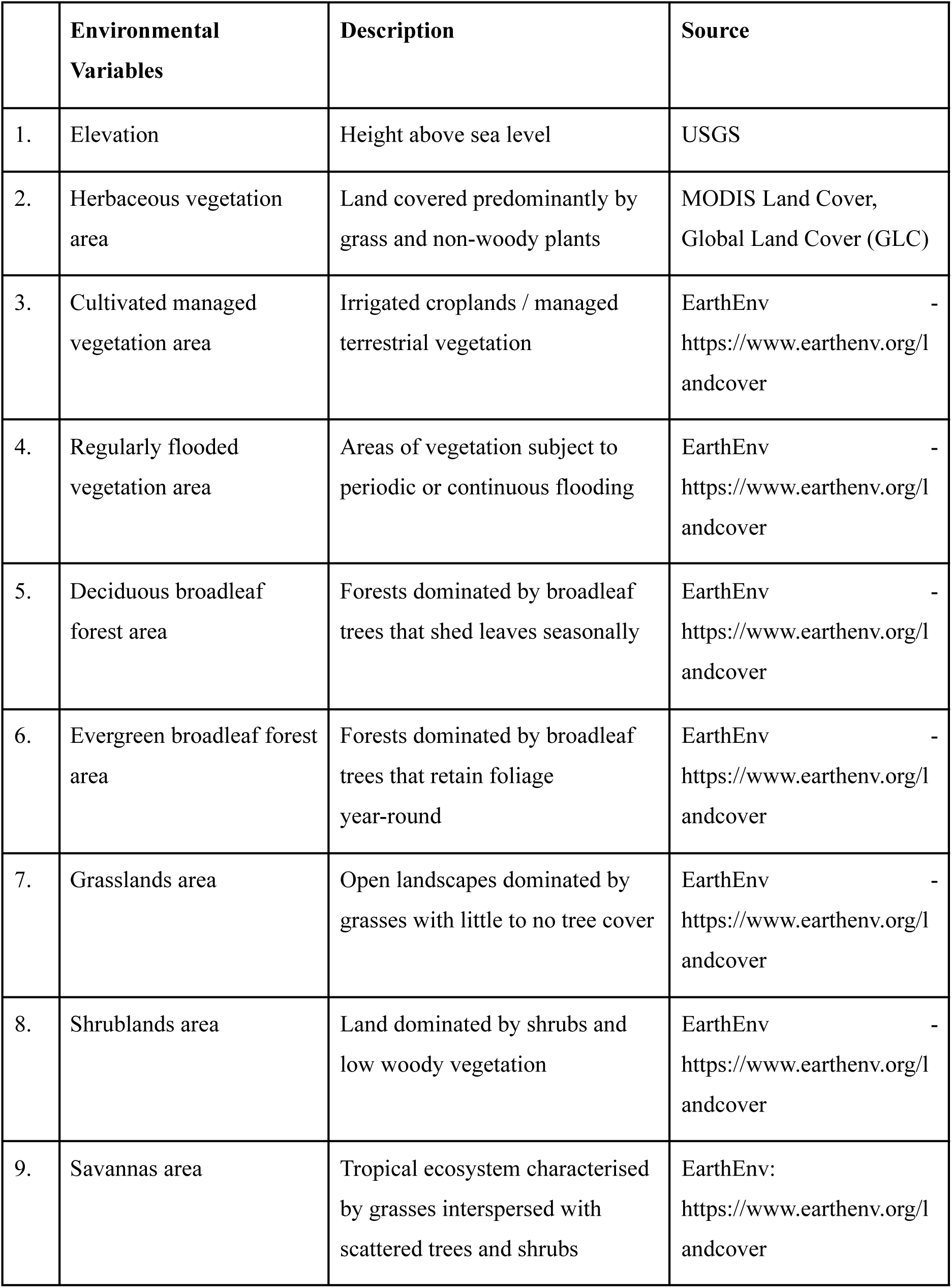
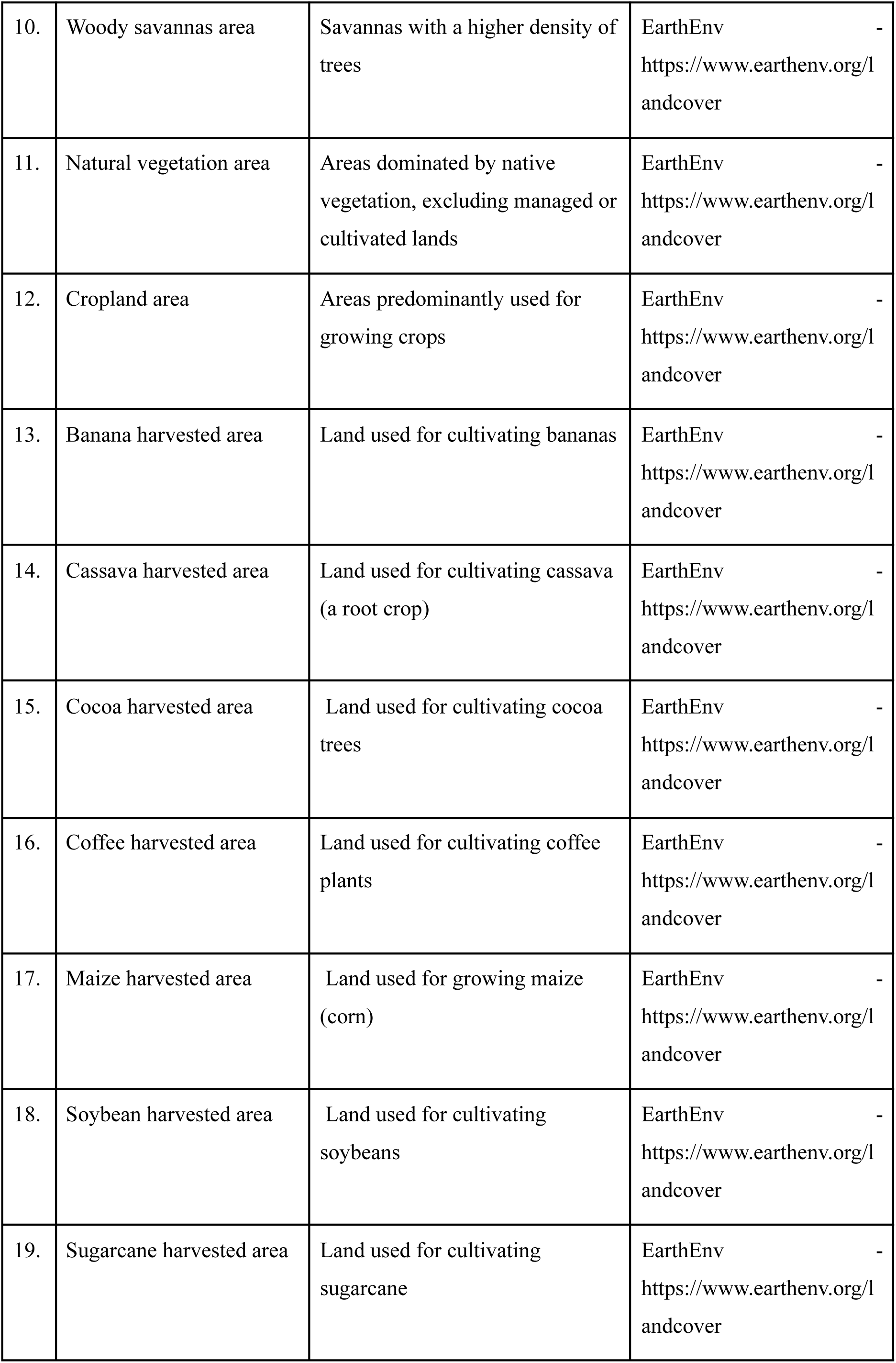
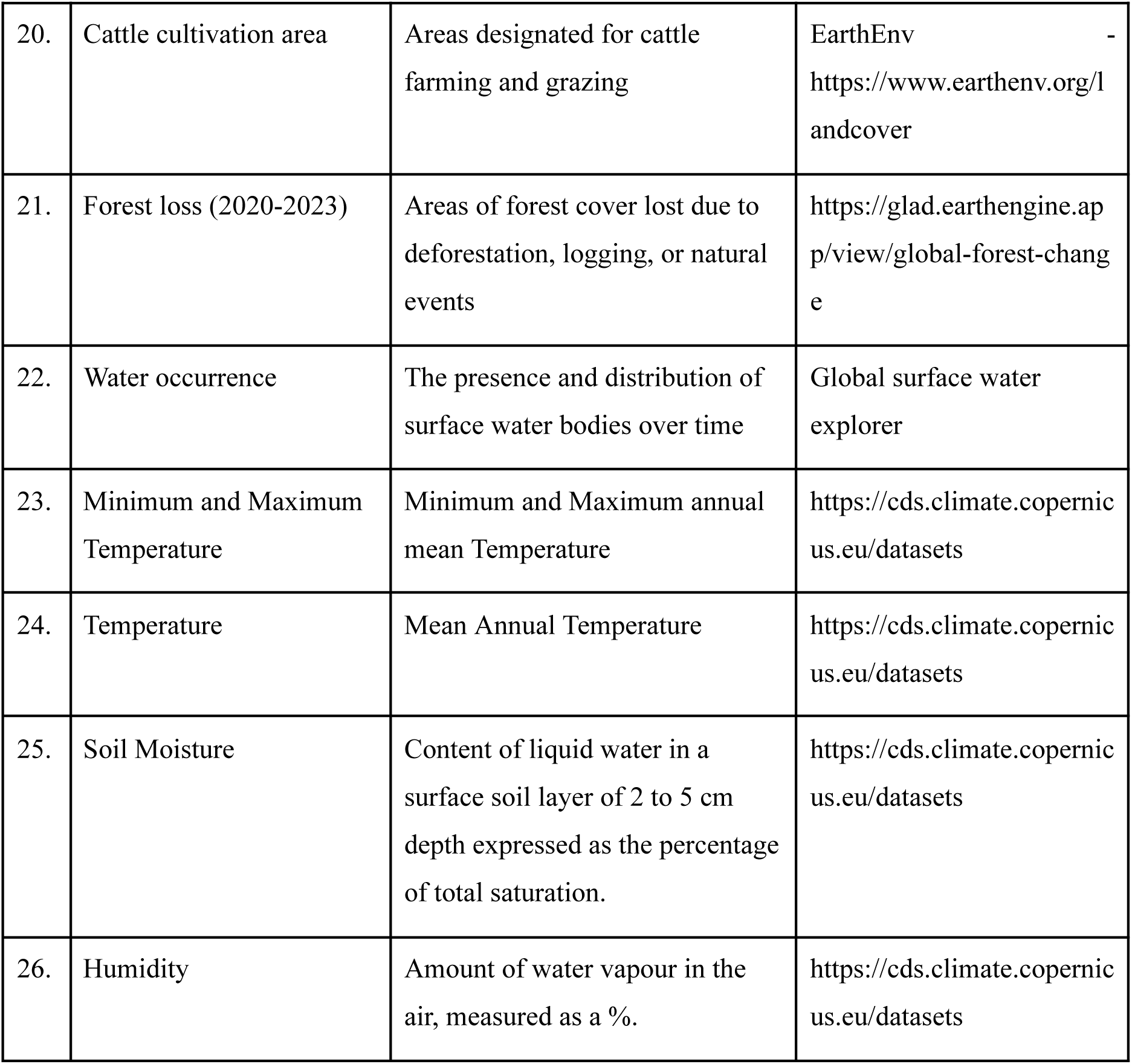
Environmental variables used in the study with respective description and data sources.

## Notes

### Competing Interest Statement

The authors have declared no competing interest.

### Summary of Updates

The Results section now includes additional validation of predictions across Central and South America, along with a SHAP analysis of the environmental factors influencing OROV distribution in Brazil. Figure 1,3 and 4 have been updated.

