## Supplementary Table S1 for "Spatiotemporal disease suitability prediction for Oropouche virus and the role of vectors across the Americas"

**Table S1: Environmental variables used in the study with respective description and data sources.**

| | **Environmental Variables** | **Description** | **Source** |
|---|---|---|---|
| 1. | Elevation | Height above sea level | USGS |
| 2. | Herbaceous vegetation area | Land covered predominantly by grass and non-woody plants | MODIS Land Cover, Global Land Cover (GLC) |
| 3. | Cultivated managed vegetation area | Irrigated croplands / managed terrestrial vegetation | EarthEnv - https://www.earthenv.org/landcover |
| 4. | Regularly flooded vegetation area | Areas of vegetation subject to periodic or continuous flooding | EarthEnv - https://www.earthenv.org/landcover |
| 5. | Deciduous broadleaf forest area | Forests dominated by broadleaf trees that shed leaves seasonally | EarthEnv - https://www.earthenv.org/landcover |
| 6. | Evergreen broadleaf forest area | Forests dominated by broadleaf trees that retain foliage year-round | EarthEnv - https://www.earthenv.org/landcover |
| 7. | Grasslands area | Open landscapes dominated by grasses with little to no tree cover | EarthEnv - https://www.earthenv.org/landcover |
| 8. | Shrublands area | Land dominated by shrubs and low woody vegetation | EarthEnv - https://www.earthenv.org/landcover |
| 9. | Savannas area | Tropical ecosystem characterised by grasses interspersed with scattered trees and shrubs | EarthEnv: https://www.earthenv.org/landcover |

| | | | |
|---|---|---|---|
| 10. | Woody savannas area | Savannas with a higher density of trees | EarthEnv - https://www.earthenv.org/landcover |
| 11. | Natural vegetation area | Areas dominated by native vegetation, excluding managed or cultivated lands | EarthEnv - https://www.earthenv.org/landcover |
| 12. | Cropland area | Areas predominantly used for growing crops | EarthEnv - https://www.earthenv.org/landcover |
| 13. | Banana harvested area | Land used for cultivating bananas | EarthEnv - https://www.earthenv.org/landcover |
| 14. | Cassava harvested area | Land used for cultivating cassava (a root crop) | EarthEnv - https://www.earthenv.org/landcover |
| 15. | Cocoa harvested area | Land used for cultivating cocoa trees | EarthEnv - https://www.earthenv.org/landcover |
| 16. | Coffee harvested area | Land used for cultivating coffee plants | EarthEnv - https://www.earthenv.org/landcover |
| 17. | Maize harvested area | Land used for growing maize (corn) | EarthEnv - https://www.earthenv.org/landcover |
| 18. | Soybean harvested area | Land used for cultivating soybeans | EarthEnv - https://www.earthenv.org/landcover |
| 19. | Sugarcane harvested area | Land used for cultivating sugarcane | EarthEnv - https://www.earthenv.org/landcover |

| | | | |
|---|---|---|---|
| 20. | Cattle cultivation area | Areas designated for cattle farming and grazing | EarthEnv - https://www.earthenv.org/landcover |
| 21. | Forest loss (2020-2023) | Areas of forest cover lost due to deforestation, logging, or natural events | https://glad.earthengine.app/view/global-forest-change |
| 22. | Water occurrence | The presence and distribution of surface water bodies over time | Global surface water explorer |
| 23. | Minimum and Maximum Temperature | Minimum and Maximum annual mean Temperature | https://cds.climate.copernicus.eu/datasets |
| 24. | Temperature | Mean Annual Temperature | https://cds.climate.copernicus.eu/datasets |
| 25. | Soil Moisture | Content of liquid water in a surface soil layer of 2 to 5 cm depth expressed as the percentage of total saturation. | https://cds.climate.copernicus.eu/datasets |
| 26. | Humidity | Amount of water vapour in the air, measured as a %. | https://cds.climate.copernicus.eu/datasets |
