## Supplementary Annexe 1 for "Spatiotemporal disease suitability prediction for Oropouche virus and the role of vectors across the Americas"

| Sampling Technique | Sigma | Radii | PA Ratio | Evaluation Metric | MeanVal | min | max |
|---|---|---|---|---|---|---|---|
| Target Group Based Approach | 1 | 0 | 400 | ROC | 0.81566667 | 0.776 | 0.945 |
| Target Group Based Approach | 1 | 0 | 400 | TSS | 0.46266667 | 0.258 | 0.568 |
| Target Group Based Approach | 1 | 0 | 800 | ROC | 0.81316667 | 0.774 | 0.904 |
| Target Group Based Approach | 1 | 0 | 800 | TSS | 0.50241667 | 0.399 | 0.656 |
| Target Group Based Approach | 1 | 0 | 2000 | ROC | 0.79525 | 0.745 | 0.866 |
| Target Group Based Approach | 1 | 0 | 2000 | TSS | 0.4215 | 0.176 | 0.611 |
| Target Group Based Approach | 1 | 0 | 4000 | ROC | 0.78883333 | 0.728 | 0.873 |
| Target Group Based Approach | 1 | 0 | 4000 | TSS | 0.35666667 | 0.163 | 0.583 |
| Target Group Based Approach | 1 | 0 | 10000 | ROC | 0.76416667 | 0.644 | 0.851 |
| Target Group Based Approach | 1 | 0 | 10000 | TSS | 0.23675 | 0.109 | 0.437 |
| Target Group Based Approach | 1 | 50-150 km | 400 | ROC | 0.85133333 | 0.747 | 0.949 |
| Target Group Based Approach | 1 | 50-150 km | 400 | TSS | 0.47183333 | 0.132 | 0.805 |
| Target Group Based Approach | 1 | 50-150 km | 800 | ROC | 0.85916667 | 0.778 | 0.933 |
| Target Group Based Approach | 1 | 50-150 km | 800 | TSS | 0.52458333 | 0.216 | 0.842 |
| Target Group Based Approach | 1 | 50-150 km | 2000 | ROC | 0.85125 | 0.745 | 0.939 |
| Target Group Based Approach | 1 | 50-150 km | 2000 | TSS | 0.44858333 | 0.129 | 0.726 |
| Target Group Based Approach | 1 | 50-150 km | 4000 | ROC | 0.83175 | 0.69 | 0.939 |
| Target Group Based Approach | 1 | 50-150 km | 4000 | TSS | 0.4225 | 0.086 | 0.666 |
| Target Group Based Approach | 1 | 50-150 km | 10000 | ROC | 0.81191667 | 0.633 | 0.91 |
| Target Group Based Approach | 1 | 50-150 km | 10000 | TSS | 0.38633333 | 0.084 | 0.63 |
| Target Group Based Approach | 1 | 50-300 km | 400 | ROC | 0.86583333 | 0.801 | 0.944 |
| Target Group Based Approach | 1 | 50-300 km | 400 | TSS | 0.555 | 0.281 | 0.745 |
| Target Group Based Approach | 1 | 50-300 km | 800 | ROC | 0.87783333 | 0.798 | 0.95 |
| Target Group Based Approach | 1 | 50-300 km | 800 | TSS | 0.56741667 | 0.303 | 0.758 |
| Target Group Based Approach | 1 | 50-300 km | 2000 | ROC | 0.86008333 | 0.769 | 0.946 |
| Target Group Based Approach | 1 | 50-300 km | 2000 | TSS | 0.49191667 | 0.243 | 0.726 |
| Target Group Based Approach | 1 | 50-300 km | 4000 | ROC | 0.85058333 | 0.762 | 0.918 |

| | | | | | | | |
|---|---|---|---|---|---|---|---|
| Target Group Based Approach | 1 | 50-300 km | 4000 | TSS | 0.46141667 | 0.23 | 0.66 |
| Target Group Based Approach | 1 | 50-300 km | 10000 | ROC | 0.82491667 | 0.693 | 0.906 |
| Target Group Based Approach | 1 | 50-300 km | 10000 | TSS | 0.4085 | 0.125 | 0.648 |
| Target Group Based Approach | 1 | 50-500km | 400 | ROC | 0.85383333 | 0.775 | 0.97 |
| Target Group Based Approach | 1 | 50-500km | 400 | TSS | 0.55333333 | 0.244 | 0.791 |
| Target Group Based Approach | 1 | 50-500km | 800 | ROC | 0.87425 | 0.784 | 0.945 |
| Target Group Based Approach | 1 | 50-500km | 800 | TSS | 0.59783333 | 0.326 | 0.793 |
| Target Group Based Approach | 1 | 50-500km | 2000 | ROC | 0.86866667 | 0.792 | 0.922 |
| Target Group Based Approach | 1 | 50-500km | 2000 | TSS | 0.53508333 | 0.343 | 0.782 |
| Target Group Based Approach | 1 | 50-500km | 4000 | ROC | 0.85616667 | 0.763 | 0.925 |
| Target Group Based Approach | 1 | 50-500km | 4000 | TSS | 0.49875 | 0.266 | 0.688 |
| Target Group Based Approach | 1 | 50-500km | 10000 | ROC | 0.8435 | 0.728 | 0.931 |
| Target Group Based Approach | 1 | 50-500km | 10000 | TSS | 0.42516667 | 0.158 | 0.649 |
| Target Group Based Approach | 3 | 0 | 400 | ROC | 0.84283333 | 0.79 | 0.932 |
| Target Group Based Approach | 3 | 0 | 400 | TSS | 0.55375 | 0.356 | 0.839 |
| Target Group Based Approach | 3 | 0 | 800 | ROC | 0.827 | 0.762 | 0.928 |
| Target Group Based Approach | 3 | 0 | 800 | TSS | 0.49991667 | 0.33 | 0.775 |
| Target Group Based Approach | 3 | 0 | 2000 | ROC | 0.82475 | 0.767 | 0.906 |
| Target Group Based Approach | 3 | 0 | 2000 | TSS | 0.44558333 | 0.261 | 0.736 |
| Target Group Based Approach | 3 | 0 | 4000 | ROC | 0.82333333 | 0.77 | 0.93 |
| Target Group Based Approach | 3 | 0 | 4000 | TSS | 0.44333333 | 0.195 | 0.652 |
| Target Group Based Approach | 3 | 0 | 10000 | ROC | 0.80025 | 0.704 | 0.899 |
| Target Group Based Approach | 3 | 0 | 10000 | TSS | 0.29983333 | 0.103 | 0.446 |
| Target Group Based Approach | 3 | 50-150 km | 400 | ROC | 0.875 | 0.756 | 0.96 |
| Target Group Based Approach | 3 | 50-150 km | 400 | TSS | 0.47316667 | 0.114 | 0.843 |
| Target Group Based Approach | 3 | 50-150 km | 800 | ROC | 0.86875 | 0.775 | 0.965 |
| Target Group Based Approach | 3 | 50-150 km | 800 | TSS | 0.54575 | 0.289 | 0.742 |
| Target Group Based Approach | 3 | 50-150 km | 2000 | ROC | 0.85591667 | 0.721 | 0.95 |

| | | | | | | | |
|---|---|---|---|---|---|---|---|
| Target Group Based Approach | 3 | 50-150 km | 2000 | TSS | 0.47375 | 0.121 | 0.783 |
| Target Group Based Approach | 3 | 50-150 km | 4000 | ROC | 0.83483333 | 0.685 | 0.948 |
| Target Group Based Approach | 3 | 50-150 km | 4000 | TSS | 0.431 | 0.14 | 0.651 |
| Target Group Based Approach | 3 | 50-150 km | 10000 | ROC | 0.81483333 | 0.647 | 0.924 |
| Target Group Based Approach | 3 | 50-150 km | 10000 | TSS | 0.37741667 | 0.107 | 0.625 |
| Target Group Based Approach | 3 | 50-300 km | 400 | ROC | 0.865 | 0.79 | 0.965 |
| Target Group Based Approach | 3 | 50-300 km | 400 | TSS | 0.53333333 | 0.266 | 0.883 |
| Target Group Based Approach | 3 | 50-300 km | 800 | ROC | 0.89433333 | 0.811 | 0.956 |
| Target Group Based Approach | 3 | 50-300 km | 800 | TSS | 0.62283333 | 0.38 | 0.851 |
| Target Group Based Approach | 3 | 50-300 km | 2000 | ROC | 0.88075 | 0.8 | 0.95 |
| Target Group Based Approach | 3 | 50-300 km | 2000 | TSS | 0.558 | 0.226 | 0.838 |
| Target Group Based Approach | 3 | 50-300 km | 4000 | ROC | 0.8715 | 0.792 | 0.944 |
| Target Group Based Approach | 3 | 50-300 km | 4000 | TSS | 0.51558333 | 0.308 | 0.651 |
| Target Group Based Approach | 3 | 50-300 km | 10000 | ROC | 0.855 | 0.739 | 0.935 |
| Target Group Based Approach | 3 | 50-300 km | 10000 | TSS | 0.45741667 | 0.197 | 0.69 |
| Target Group Based Approach | 3 | 50-500km | 400 | ROC | 0.88675 | 0.808 | 0.976 |
| Target Group Based Approach | 3 | 50-500km | 400 | TSS | 0.61833333 | 0.339 | 0.945 |
| Target Group Based Approach | 3 | 50-500km | 800 | ROC | 0.89408333 | 0.827 | 0.953 |
| Target Group Based Approach | 3 | 50-500km | 800 | TSS | 0.65025 | 0.433 | 0.846 |
| Target Group Based Approach | 3 | 50-500km | 2000 | ROC | 0.88141667 | 0.804 | 0.937 |
| Target Group Based Approach | 3 | 50-500km | 2000 | TSS | 0.59983333 | 0.459 | 0.777 |
| Target Group Based Approach | 3 | 50-500km | 4000 | ROC | 0.87533333 | 0.797 | 0.944 |
| Target Group Based Approach | 3 | 50-500km | 4000 | TSS | 0.51741667 | 0.294 | 0.715 |
| Target Group Based Approach | 3 | 50-500km | 10000 | ROC | 0.85516667 | 0.753 | 0.933 |
| Target Group Based Approach | 3 | 50-500km | 10000 | TSS | 0.46808333 | 0.253 | 0.679 |
| Target Group Based Approach | 5 | 0 | 400 | ROC | 0.82175 | 0.729 | 0.915 |
| Target Group Based Approach | 5 | 0 | 400 | TSS | 0.48766667 | 0.229 | 0.684 |
| Target Group Based Approach | 5 | 0 | 800 | ROC | 0.84475 | 0.791 | 0.94 |

| | | | | | | | |
|---|---|---|---|---|---|---|---|
| Target Group Based Approach | 5 | 0 | 800 | TSS | 0.53408333 | 0.274 | 0.777 |
| Target Group Based Approach | 5 | 0 | 2000 | ROC | 0.84233333 | 0.777 | 0.925 |
| Target Group Based Approach | 5 | 0 | 2000 | TSS | 0.50608333 | 0.157 | 0.734 |
| Target Group Based Approach | 5 | 0 | 4000 | ROC | 0.82225 | 0.76 | 0.91 |
| Target Group Based Approach | 5 | 0 | 4000 | TSS | 0.40441667 | 0.157 | 0.508 |
| Target Group Based Approach | 5 | 0 | 10000 | ROC | 0.80525 | 0.717 | 0.904 |
| Target Group Based Approach | 5 | 0 | 10000 | TSS | 0.25516667 | 0.097 | 0.498 |
| Target Group Based Approach | 5 | 50-150 km | 400 | ROC | 0.857 | 0.749 | 0.953 |
| Target Group Based Approach | 5 | 50-150 km | 400 | TSS | 0.51458333 | 0.088 | 0.802 |
| Target Group Based Approach | 5 | 50-150 km | 800 | ROC | 0.87308333 | 0.773 | 0.958 |
| Target Group Based Approach | 5 | 50-150 km | 800 | TSS | 0.5565 | 0.273 | 0.812 |
| Target Group Based Approach | 5 | 50-150 km | 2000 | ROC | 0.85225 | 0.739 | 0.947 |
| Target Group Based Approach | 5 | 50-150 km | 2000 | TSS | 0.45758333 | 0.164 | 0.741 |
| Target Group Based Approach | 5 | 50-150 km | 4000 | ROC | 0.82841667 | 0.675 | 0.928 |
| Target Group Based Approach | 5 | 50-150 km | 4000 | TSS | 0.42708333 | 0.11 | 0.635 |
| Target Group Based Approach | 5 | 50-150 km | 10000 | ROC | 0.80291667 | 0.605 | 0.909 |
| Target Group Based Approach | 5 | 50-150 km | 10000 | TSS | 0.38066667 | 0.076 | 0.623 |
| Target Group Based Approach | 5 | 50-300 km | 400 | ROC | 0.886 | 0.81 | 0.963 |
| Target Group Based Approach | 5 | 50-300 km | 400 | TSS | 0.59466667 | 0.466 | 0.843 |
| Target Group Based Approach | 5 | 50-300 km | 800 | ROC | 0.89283333 | 0.824 | 0.959 |
| Target Group Based Approach | 5 | 50-300 km | 800 | TSS | 0.61566667 | 0.422 | 0.826 |
| Target Group Based Approach | 5 | 50-300 km | 2000 | ROC | 0.88375 | 0.807 | 0.944 |
| Target Group Based Approach | 5 | 50-300 km | 2000 | TSS | 0.56283333 | 0.286 | 0.767 |
| Target Group Based Approach | 5 | 50-300 km | 4000 | ROC | 0.869 | 0.796 | 0.948 |
| Target Group Based Approach | 5 | 50-300 km | 4000 | TSS | 0.52241667 | 0.27 | 0.675 |
| Target Group Based Approach | 5 | 50-300 km | 10000 | ROC | 0.85341667 | 0.747 | 0.931 |
| Target Group Based Approach | 5 | 50-300 km | 10000 | TSS | 0.47825 | 0.257 | 0.673 |
| Target Group Based Approach | 5 | 50-500km | 400 | ROC | 0.90183333 | 0.843 | 0.965 |

| | | | | | | | |
|---|---|---|---|---|---|---|---|
| Target Group Based Approach | | 5 | 50-500km | 400 | TSS | 0.74558333 | 0.669 | 0.809 |
| Target Group Based Approach | | 5 | 50-500km | 800 | ROC | 0.90575 | 0.836 | 0.961 |
| Target Group Based Approach | | 5 | 50-500km | 800 | TSS | 0.64725 | 0.506 | 0.782 |
| Target Group Based Approach | | 5 | 50-500km | 2000 | ROC | 0.88466667 | 0.822 | 0.952 |
| Target Group Based Approach | | 5 | 50-500km | 2000 | TSS | 0.56675 | 0.323 | 0.81 |
| Target Group Based Approach | | 5 | 50-500km | 4000 | ROC | 0.88483333 | 0.808 | 0.953 |
| Target Group Based Approach | | 5 | 50-500km | 4000 | TSS | 0.568 | 0.349 | 0.668 |
| Target Group Based Approach | | 5 | 50-500km | 10000 | ROC | 0.85741667 | 0.77 | 0.937 |
| Target Group Based Approach | | 5 | 50-500km | 10000 | TSS | 0.48116667 | 0.286 | 0.655 |
| Random Sampling | - | | - | 400 | ROC | 0.83891667 | 0.73 | 0.938 |
| Random Sampling | - | | - | 400 | TSS | 0.49783333 | 0.164 | 0.76 |
| Random Sampling | - | | - | 800 | ROC | 0.85275 | 0.78 | 0.94 |
| Random Sampling | - | | - | 800 | TSS | 0.53791667 | 0.412 | 0.741 |
| Random Sampling | - | | - | 2000 | ROC | 0.83916667 | 0.745 | 0.925 |
| Random Sampling | - | | - | 2000 | TSS | 0.45583333 | 0.161 | 0.59 |
| Random Sampling | - | | - | 4000 | ROC | 0.82141667 | 0.746 | 0.907 |
| Random Sampling | - | | - | 4000 | TSS | 0.33258333 | 0.159 | 0.532 |
| Random Sampling | - | | - | 10000 | ROC | 0.60366667 | 0.509 | 0.749 |
| Random Sampling | - | | - | 10000 | TSS | 0.258 | 0.042 | 0.506 |
| Density-Weighted Population-Based Approach | | 1 | 0 | 400 | ROC | 0.68191667 | 0.646 | 0.731 |
| Density-Weighted Population-Based Approach | | 1 | 0 | 400 | TSS | 0.25916667 | 0.187 | 0.354 |
| Density-Weighted Population-Based Approach | | 1 | 0 | 800 | ROC | 0.66991667 | 0.545 | 0.81 |
| Density-Weighted Population-Based Approach | | 1 | 0 | 800 | TSS | 0.21641667 | 0.081 | 0.422 |
| Density-Weighted Population-Based Approach | | 1 | 0 | 2000 | ROC | 0.65533333 | 0.574 | 0.773 |
| Density-Weighted Population-Based Approach | | 1 | 0 | 2000 | TSS | 0.19383333 | 0.058 | 0.386 |
| Density-Weighted Population-Based Approach | | 1 | 0 | 4000 | ROC | 0.64641667 | 0.513 | 0.765 |
| Density-Weighted Population-Based Approach | | 1 | 0 | 4000 | TSS | 0.14716667 | 0.004 | 0.277 |
| Density-Weighted Population-Based Approach | | 1 | 0 | 10000 | ROC | 0.63666667 | 0.473 | 0.797 |

| | | | | | | | |
|---|---|---|---|---|---|---|---|
| Density-Weighted Population-Based Approach | 1 | 0 | 10000 | TSS | 0.07491667 | -0.031 | 0.158 |
| Density-Weighted Population-Based Approach | 1 | 50-150 km | 400 | ROC | 0.80625 | 0.682 | 0.933 |
| Density-Weighted Population-Based Approach | 1 | 50-150 km | 400 | TSS | 0.35825 | 0.159 | 0.661 |
| Density-Weighted Population-Based Approach | 1 | 50-150 km | 800 | ROC | 0.80758333 | 0.692 | 0.928 |
| Density-Weighted Population-Based Approach | 1 | 50-150 km | 800 | TSS | 0.38525 | 0.163 | 0.715 |
| Density-Weighted Population-Based Approach | 1 | 50-150 km | 2000 | ROC | 0.79133333 | 0.634 | 0.905 |
| Density-Weighted Population-Based Approach | 1 | 50-150 km | 2000 | TSS | 0.37491667 | 0.135 | 0.658 |
| Density-Weighted Population-Based Approach | 1 | 50-150 km | 4000 | ROC | 0.76583333 | 0.598 | 0.877 |
| Density-Weighted Population-Based Approach | 1 | 50-150 km | 4000 | TSS | 0.34575 | 0.088 | 0.61 |
| Density-Weighted Population-Based Approach | 1 | 50-150 km | 10000 | ROC | 0.74 | 0.56 | 0.856 |
| Density-Weighted Population-Based Approach | 1 | 50-150 km | 10000 | TSS | 0.28975 | 0.059 | 0.596 |
| Density-Weighted Population-Based Approach | 1 | 50-300 km | 400 | ROC | 0.82458333 | 0.753 | 0.94 |
| Density-Weighted Population-Based Approach | 1 | 50-300 km | 400 | TSS | 0.4405 | 0.247 | 0.707 |
| Density-Weighted Population-Based Approach | 1 | 50-300 km | 800 | ROC | 0.82083333 | 0.686 | 0.923 |
| Density-Weighted Population-Based Approach | 1 | 50-300 km | 800 | TSS | 0.40083333 | 0.138 | 0.703 |
| Density-Weighted Population-Based Approach | 1 | 50-300 km | 2000 | ROC | 0.79616667 | 0.632 | 0.907 |
| Density-Weighted Population-Based Approach | 1 | 50-300 km | 2000 | TSS | 0.38441667 | 0.152 | 0.667 |
| Density-Weighted Population-Based Approach | 1 | 50-300 km | 4000 | ROC | 0.76858333 | 0.601 | 0.862 |
| Density-Weighted Population-Based Approach | 1 | 50-300 km | 4000 | TSS | 0.34841667 | 0.097 | 0.565 |
| Density-Weighted Population-Based Approach | 1 | 50-300 km | 10000 | ROC | 0.7505 | 0.565 | 0.888 |
| Density-Weighted Population-Based Approach | 1 | 50-300 km | 10000 | TSS | 0.30608333 | 0.079 | 0.607 |
| Density-Weighted Population-Based Approach | 1 | 50-500km | 400 | ROC | 0.80275 | 0.699 | 0.894 |
| Density-Weighted Population-Based Approach | 1 | 50-500km | 400 | TSS | 0.38258333 | 0.223 | 0.673 |
| Density-Weighted Population-Based Approach | 1 | 50-500km | 800 | ROC | 0.82233333 | 0.715 | 0.915 |
| Density-Weighted Population-Based Approach | 1 | 50-500km | 800 | TSS | 0.45408333 | 0.194 | 0.729 |
| Density-Weighted Population-Based Approach | 1 | 50-500km | 2000 | ROC | 0.79425 | 0.638 | 0.905 |
| Density-Weighted Population-Based Approach | 1 | 50-500km | 2000 | TSS | 0.38333333 | 0.097 | 0.702 |
| Density-Weighted Population-Based Approach | 1 | 50-500km | 4000 | ROC | 0.76833333 | 0.607 | 0.882 |

| | | | | | | | |
|---|---|---|---|---|---|---|---|
| Density-Weighted Population-Based Approach | 1 | 50-500km | 4000 | TSS | 0.35575 | 0.088 | 0.627 |
| Density-Weighted Population-Based Approach | 1 | 50-500km | 10000 | ROC | 0.7495 | 0.56 | 0.871 |
| Density-Weighted Population-Based Approach | 1 | 50-500km | 10000 | TSS | 0.29958333 | 0.082 | 0.61 |
| Density-Weighted Population-Based Approach | 3 | 0 | 400 | ROC | 0.742 | 0.686 | 0.796 |
| Density-Weighted Population-Based Approach | 3 | 0 | 400 | TSS | 0.33333333 | 0.184 | 0.486 |
| Density-Weighted Population-Based Approach | 3 | 0 | 800 | ROC | 0.76625 | 0.722 | 0.837 |
| Density-Weighted Population-Based Approach | 3 | 0 | 800 | TSS | 0.384 | 0.259 | 0.579 |
| Density-Weighted Population-Based Approach | 3 | 0 | 2000 | ROC | 0.735 | 0.689 | 0.767 |
| Density-Weighted Population-Based Approach | 3 | 0 | 2000 | TSS | 0.3145 | 0.216 | 0.434 |
| Density-Weighted Population-Based Approach | 3 | 0 | 4000 | ROC | 0.72425 | 0.621 | 0.783 |
| Density-Weighted Population-Based Approach | 3 | 0 | 4000 | TSS | 0.22966667 | 0.132 | 0.367 |
| Density-Weighted Population-Based Approach | 3 | 0 | 10000 | ROC | 0.69191667 | 0.564 | 0.761 |
| Density-Weighted Population-Based Approach | 3 | 0 | 10000 | TSS | 0.13783333 | -0.003 | 0.3 |
| Density-Weighted Population-Based Approach | 3 | 50-150 km | 400 | ROC | 0.8165 | 0.733 | 0.923 |
| Density-Weighted Population-Based Approach | 3 | 50-150 km | 400 | TSS | 0.44475 | 0.223 | 0.753 |
| Density-Weighted Population-Based Approach | 3 | 50-150 km | 800 | ROC | 0.82091667 | 0.723 | 0.913 |
| Density-Weighted Population-Based Approach | 3 | 50-150 km | 800 | TSS | 0.44283333 | 0.16 | 0.686 |
| Density-Weighted Population-Based Approach | 3 | 50-150 km | 2000 | ROC | 0.8115 | 0.68 | 0.895 |
| Density-Weighted Population-Based Approach | 3 | 50-150 km | 2000 | TSS | 0.38183333 | 0.181 | 0.657 |
| Density-Weighted Population-Based Approach | 3 | 50-150 km | 4000 | ROC | 0.79358333 | 0.63 | 0.891 |
| Density-Weighted Population-Based Approach | 3 | 50-150 km | 4000 | TSS | 0.37791667 | 0.128 | 0.633 |
| Density-Weighted Population-Based Approach | 3 | 50-150 km | 10000 | ROC | 0.76358333 | 0.586 | 0.878 |
| Density-Weighted Population-Based Approach | 3 | 50-150 km | 10000 | TSS | 0.32858333 | 0.117 | 0.618 |
| Density-Weighted Population-Based Approach | 3 | 50-300 km | 400 | ROC | 0.84641667 | 0.76 | 0.935 |
| Density-Weighted Population-Based Approach | 3 | 50-300 km | 400 | TSS | 0.52808333 | 0.415 | 0.711 |
| Density-Weighted Population-Based Approach | 3 | 50-300 km | 800 | ROC | 0.84608333 | 0.782 | 0.916 |
| Density-Weighted Population-Based Approach | 3 | 50-300 km | 800 | TSS | 0.4905 | 0.234 | 0.721 |
| Density-Weighted Population-Based Approach | 3 | 50-300 km | 2000 | ROC | 0.8315 | 0.731 | 0.907 |

| | | | | | | | |
|---|---|---|---|---|---|---|---|
| Density-Weighted Population-Based Approach | 3 | 50-300 km | 2000 | TSS | 0.42158333 | 0.118 | 0.64 |
| Density-Weighted Population-Based Approach | 3 | 50-300 km | 4000 | ROC | 0.81625 | 0.692 | 0.906 |
| Density-Weighted Population-Based Approach | 3 | 50-300 km | 4000 | TSS | 0.41941667 | 0.146 | 0.623 |
| Density-Weighted Population-Based Approach | 3 | 50-300 km | 10000 | ROC | 0.80225 | 0.647 | 0.903 |
| Density-Weighted Population-Based Approach | 3 | 50-300 km | 10000 | TSS | 0.36783333 | 0.139 | 0.62 |
| Density-Weighted Population-Based Approach | 3 | 50-500km | 400 | ROC | 0.83275 | 0.746 | 0.934 |
| Density-Weighted Population-Based Approach | 3 | 50-500km | 400 | TSS | 0.42858333 | -0.011 | 0.664 |
| Density-Weighted Population-Based Approach | 3 | 50-500km | 800 | ROC | 0.85808333 | 0.771 | 0.937 |
| Density-Weighted Population-Based Approach | 3 | 50-500km | 800 | TSS | 0.51675 | 0.335 | 0.746 |
| Density-Weighted Population-Based Approach | 3 | 50-500km | 2000 | ROC | 0.84141667 | 0.763 | 0.909 |
| Density-Weighted Population-Based Approach | 3 | 50-500km | 2000 | TSS | 0.48041667 | 0.268 | 0.655 |
| Density-Weighted Population-Based Approach | 3 | 50-500km | 4000 | ROC | 0.82058333 | 0.712 | 0.903 |
| Density-Weighted Population-Based Approach | 3 | 50-500km | 4000 | TSS | 0.42483333 | 0.22 | 0.647 |
| Density-Weighted Population-Based Approach | 3 | 50-500km | 10000 | ROC | 0.80408333 | 0.655 | 0.893 |
| Density-Weighted Population-Based Approach | 3 | 50-500km | 10000 | TSS | 0.3845 | 0.169 | 0.641 |
| Density-Weighted Population-Based Approach | 5 | 0 | 400 | ROC | 0.768 | 0.731 | 0.842 |
| Density-Weighted Population-Based Approach | 5 | 0 | 400 | TSS | 0.36758333 | 0.172 | 0.511 |
| Density-Weighted Population-Based Approach | 5 | 0 | 800 | ROC | 0.74591667 | 0.538 | 0.799 |
| Density-Weighted Population-Based Approach | 5 | 0 | 800 | TSS | 0.33016667 | 0.083 | 0.465 |
| Density-Weighted Population-Based Approach | 5 | 0 | 2000 | ROC | 0.73875 | 0.696 | 0.77 |
| Density-Weighted Population-Based Approach | 5 | 0 | 2000 | TSS | 0.28641667 | 0.034 | 0.451 |
| Density-Weighted Population-Based Approach | 5 | 0 | 4000 | ROC | 0.7325 | 0.699 | 0.809 |
| Density-Weighted Population-Based Approach | 5 | 0 | 4000 | TSS | 0.24175 | 0.168 | 0.335 |
| Density-Weighted Population-Based Approach | 5 | 0 | 10000 | ROC | 0.70475 | 0.575 | 0.779 |
| Density-Weighted Population-Based Approach | 5 | 0 | 10000 | TSS | 0.13766667 | -0.002 | 0.239 |
| Density-Weighted Population-Based Approach | 5 | 50-150 km | 400 | ROC | 0.84541667 | 0.741 | 0.946 |
| Density-Weighted Population-Based Approach | 5 | 50-150 km | 400 | TSS | 0.49325 | 0.241 | 0.714 |
| Density-Weighted Population-Based Approach | 5 | 50-150 km | 800 | ROC | 0.82441667 | 0.715 | 0.908 |

| | | | | | | | |
|---|---|---|---|---|---|---|---|
| Density-Weighted Population-Based Approach | 5 | 50-150 km | 800 | TSS | 0.42675 | 0.208 | 0.662 |
| Density-Weighted Population-Based Approach | 5 | 50-150 km | 2000 | ROC | 0.81075 | 0.663 | 0.92 |
| Density-Weighted Population-Based Approach | 5 | 50-150 km | 2000 | TSS | 0.37975 | 0.145 | 0.657 |
| Density-Weighted Population-Based Approach | 5 | 50-150 km | 4000 | ROC | 0.79508333 | 0.638 | 0.888 |
| Density-Weighted Population-Based Approach | 5 | 50-150 km | 4000 | TSS | 0.37558333 | 0.114 | 0.611 |
| Density-Weighted Population-Based Approach | 5 | 50-150 km | 10000 | ROC | 0.76958333 | 0.591 | 0.872 |
| Density-Weighted Population-Based Approach | 5 | 50-150 km | 10000 | TSS | 0.31175 | 0.083 | 0.603 |
| Density-Weighted Population-Based Approach | 5 | 50-300 km | 400 | ROC | 0.85366667 | 0.785 | 0.974 |
| Density-Weighted Population-Based Approach | 5 | 50-300 km | 400 | TSS | 0.49291667 | 0.247 | 0.706 |
| Density-Weighted Population-Based Approach | 5 | 50-300 km | 800 | ROC | 0.8565 | 0.795 | 0.931 |
| Density-Weighted Population-Based Approach | 5 | 50-300 km | 800 | TSS | 0.46483333 | 0.206 | 0.693 |
| Density-Weighted Population-Based Approach | 5 | 50-300 km | 2000 | ROC | 0.85066667 | 0.755 | 0.934 |
| Density-Weighted Population-Based Approach | 5 | 50-300 km | 2000 | TSS | 0.46325 | 0.21 | 0.727 |
| Density-Weighted Population-Based Approach | 5 | 50-300 km | 4000 | ROC | 0.827 | 0.71 | 0.905 |
| Density-Weighted Population-Based Approach | 5 | 50-300 km | 4000 | TSS | 0.42975 | 0.25 | 0.61 |
| Density-Weighted Population-Based Approach | 5 | 50-300 km | 10000 | ROC | 0.8045 | 0.671 | 0.897 |
| Density-Weighted Population-Based Approach | 5 | 50-300 km | 10000 | TSS | 0.38708333 | 0.171 | 0.619 |
| Density-Weighted Population-Based Approach | 5 | 50-500km | 400 | ROC | 0.8745 | 0.797 | 0.967 |
| Density-Weighted Population-Based Approach | 5 | 50-500km | 400 | TSS | 0.52566667 | 0 | 0.82 |
| Density-Weighted Population-Based Approach | 5 | 50-500km | 800 | ROC | 0.8555 | 0.779 | 0.931 |
| Density-Weighted Population-Based Approach | 5 | 50-500km | 800 | TSS | 0.53033333 | 0.192 | 0.673 |
| Density-Weighted Population-Based Approach | 5 | 50-500km | 2000 | ROC | 0.84908333 | 0.768 | 0.917 |
| Density-Weighted Population-Based Approach | 5 | 50-500km | 2000 | TSS | 0.49391667 | 0.327 | 0.676 |
| Density-Weighted Population-Based Approach | 5 | 50-500km | 4000 | ROC | 0.83075 | 0.723 | 0.916 |
| Density-Weighted Population-Based Approach | 5 | 50-500km | 4000 | TSS | 0.45958333 | 0.21 | 0.668 |
| Density-Weighted Population-Based Approach | 5 | 50-500km | 10000 | ROC | 0.81166667 | 0.691 | 0.906 |
| Density-Weighted Population-Based Approach | 5 | 50-500km | 10000 | TSS | 0.38325 | 0.18 | 0.632 |
| Density-Weighted Geographic Sampling | 1 | 0 | 400 | ROC | 0.76175 | 0.645 | 0.917 |

| | | | | | | | |
|---|---|---|---|---|---|---|---|
| Density-Weighted Geographic Sampling | 1 | 0 | 400 | TSS | 0.40183333 | 0.088 | 0.721 |
| Density-Weighted Geographic Sampling | 1 | 0 | 800 | ROC | 0.76241667 | 0.681 | 0.898 |
| Density-Weighted Geographic Sampling | 1 | 0 | 800 | TSS | 0.33225 | 0.116 | 0.651 |
| Density-Weighted Geographic Sampling | 1 | 0 | 2000 | ROC | 0.73933333 | 0.638 | 0.871 |
| Density-Weighted Geographic Sampling | 1 | 0 | 2000 | TSS | 0.27591667 | 0.114 | 0.545 |
| Density-Weighted Geographic Sampling | 1 | 0 | 4000 | ROC | 0.71016667 | 0.575 | 0.874 |
| Density-Weighted Geographic Sampling | 1 | 0 | 4000 | TSS | 0.17225 | 0 | 0.426 |
| Density-Weighted Geographic Sampling | 1 | 0 | 10000 | ROC | 0.69741667 | 0.529 | 0.877 |
| Density-Weighted Geographic Sampling | 1 | 0 | 10000 | TSS | 0.1275 | 0 | 0.39 |
| Density-Weighted Geographic Sampling | 1 | 50-150 km | 400 | ROC | 0.84716667 | 0.762 | 0.954 |
| Density-Weighted Geographic Sampling | 1 | 50-150 km | 400 | TSS | 0.46925 | 0.127 | 0.828 |
| Density-Weighted Geographic Sampling | 1 | 50-150 km | 800 | ROC | 0.85091667 | 0.773 | 0.944 |
| Density-Weighted Geographic Sampling | 1 | 50-150 km | 800 | TSS | 0.51066667 | 0.262 | 0.808 |
| Density-Weighted Geographic Sampling | 1 | 50-150 km | 2000 | ROC | 0.84283333 | 0.734 | 0.946 |
| Density-Weighted Geographic Sampling | 1 | 50-150 km | 2000 | TSS | 0.44825 | 0.194 | 0.71 |
| Density-Weighted Geographic Sampling | 1 | 50-150 km | 4000 | ROC | 0.81641667 | 0.694 | 0.938 |
| Density-Weighted Geographic Sampling | 1 | 50-150 km | 4000 | TSS | 0.42758333 | 0.169 | 0.592 |
| Density-Weighted Geographic Sampling | 1 | 50-150 km | 10000 | ROC | 0.79641667 | 0.658 | 0.931 |
| Density-Weighted Geographic Sampling | 1 | 50-150 km | 10000 | TSS | 0.34383333 | 0.106 | 0.565 |
| Density-Weighted Geographic Sampling | 1 | 50-300 km | 400 | ROC | 0.86016667 | 0.782 | 0.952 |
| Density-Weighted Geographic Sampling | 1 | 50-300 km | 400 | TSS | 0.53266667 | 0.217 | 0.808 |
| Density-Weighted Geographic Sampling | 1 | 50-300 km | 800 | ROC | 0.86116667 | 0.792 | 0.948 |
| Density-Weighted Geographic Sampling | 1 | 50-300 km | 800 | TSS | 0.53508333 | 0.306 | 0.78 |
| Density-Weighted Geographic Sampling | 1 | 50-300 km | 2000 | ROC | 0.83525 | 0.727 | 0.937 |
| Density-Weighted Geographic Sampling | 1 | 50-300 km | 2000 | TSS | 0.46216667 | 0.194 | 0.622 |
| Density-Weighted Geographic Sampling | 1 | 50-300 km | 4000 | ROC | 0.82825 | 0.683 | 0.931 |
| Density-Weighted Geographic Sampling | 1 | 50-300 km | 4000 | TSS | 0.41791667 | 0.179 | 0.609 |
| Density-Weighted Geographic Sampling | 1 | 50-300 km | 10000 | ROC | 0.81158333 | 0.681 | 0.923 |

| | | | | | | | |
|---|---|---|---|---|---|---|---|
| Density-Weighted Geographic Sampling | 1 | 50-300 km | 10000 | TSS | 0.36433333 | 0.154 | 0.594 |
| Density-Weighted Geographic Sampling | 1 | 50-500km | 400 | ROC | 0.85116667 | 0.79 | 0.95 |
| Density-Weighted Geographic Sampling | 1 | 50-500km | 400 | TSS | 0.49383333 | 0.181 | 0.831 |
| Density-Weighted Geographic Sampling | 1 | 50-500km | 800 | ROC | 0.85083333 | 0.759 | 0.944 |
| Density-Weighted Geographic Sampling | 1 | 50-500km | 800 | TSS | 0.50466667 | 0.299 | 0.826 |
| Density-Weighted Geographic Sampling | 1 | 50-500km | 2000 | ROC | 0.85191667 | 0.757 | 0.948 |
| Density-Weighted Geographic Sampling | 1 | 50-500km | 2000 | TSS | 0.48 | 0.144 | 0.727 |
| Density-Weighted Geographic Sampling | 1 | 50-500km | 4000 | ROC | 0.82541667 | 0.688 | 0.941 |
| Density-Weighted Geographic Sampling | 1 | 50-500km | 4000 | TSS | 0.41783333 | 0.133 | 0.584 |
| Density-Weighted Geographic Sampling | 1 | 50-500km | 10000 | ROC | 0.80475 | 0.689 | 0.915 |
| Density-Weighted Geographic Sampling | 1 | 50-500km | 10000 | TSS | 0.34291667 | 0.153 | 0.573 |
| Density-Weighted Geographic Sampling | 3 | 0 | 400 | ROC | 0.84225 | 0.751 | 0.934 |
| Density-Weighted Geographic Sampling | 3 | 0 | 400 | TSS | 0.51466667 | 0.245 | 0.797 |
| Density-Weighted Geographic Sampling | 3 | 0 | 800 | ROC | 0.84141667 | 0.794 | 0.918 |
| Density-Weighted Geographic Sampling | 3 | 0 | 800 | TSS | 0.49383333 | 0.43 | 0.67 |
| Density-Weighted Geographic Sampling | 3 | 0 | 2000 | ROC | 0.816 | 0.744 | 0.923 |
| Density-Weighted Geographic Sampling | 3 | 0 | 2000 | TSS | 0.34641667 | 0.198 | 0.562 |
| Density-Weighted Geographic Sampling | 3 | 0 | 4000 | ROC | 0.81191667 | 0.68 | 0.92 |
| Density-Weighted Geographic Sampling | 3 | 0 | 4000 | TSS | 0.35516667 | 0.216 | 0.518 |
| Density-Weighted Geographic Sampling | 3 | 0 | 10000 | ROC | 0.786 | 0.641 | 0.911 |
| Density-Weighted Geographic Sampling | 3 | 0 | 10000 | TSS | 0.23675 | 0.011 | 0.443 |
| Density-Weighted Geographic Sampling | 3 | 50-150 km | 400 | ROC | 0.8705 | 0.796 | 0.958 |
| Density-Weighted Geographic Sampling | 3 | 50-150 km | 400 | TSS | 0.57458333 | 0.3 | 0.863 |
| Density-Weighted Geographic Sampling | 3 | 50-150 km | 800 | ROC | 0.87033333 | 0.783 | 0.948 |
| Density-Weighted Geographic Sampling | 3 | 50-150 km | 800 | TSS | 0.55383333 | 0.294 | 0.83 |
| Density-Weighted Geographic Sampling | 3 | 50-150 km | 2000 | ROC | 0.85991667 | 0.755 | 0.949 |
| Density-Weighted Geographic Sampling | 3 | 50-150 km | 2000 | TSS | 0.51383333 | 0.256 | 0.691 |
| Density-Weighted Geographic Sampling | 3 | 50-150 km | 4000 | ROC | 0.834 | 0.69 | 0.938 |

| | | | | | | | |
|---|---|---|---|---|---|---|---|
| Density-Weighted Geographic Sampling | 3 | 50-150 km | 4000 | TSS | 0.46308333 | 0.221 | 0.651 |
| Density-Weighted Geographic Sampling | 3 | 50-150 km | 10000 | ROC | 0.808 | 0.647 | 0.923 |
| Density-Weighted Geographic Sampling | 3 | 50-150 km | 10000 | TSS | 0.37875 | 0.175 | 0.614 |
| Density-Weighted Geographic Sampling | 3 | 50-300 km | 400 | ROC | 0.894 | 0.813 | 0.959 |
| Density-Weighted Geographic Sampling | 3 | 50-300 km | 400 | TSS | 0.604 | 0.411 | 0.897 |
| Density-Weighted Geographic Sampling | 3 | 50-300 km | 800 | ROC | 0.88991667 | 0.819 | 0.949 |
| Density-Weighted Geographic Sampling | 3 | 50-300 km | 800 | TSS | 0.60325 | 0.326 | 0.809 |
| Density-Weighted Geographic Sampling | 3 | 50-300 km | 2000 | ROC | 0.878 | 0.795 | 0.946 |
| Density-Weighted Geographic Sampling | 3 | 50-300 km | 2000 | TSS | 0.53166667 | 0.28 | 0.701 |
| Density-Weighted Geographic Sampling | 3 | 50-300 km | 4000 | ROC | 0.86433333 | 0.768 | 0.945 |
| Density-Weighted Geographic Sampling | 3 | 50-300 km | 4000 | TSS | 0.49975 | 0.296 | 0.683 |
| Density-Weighted Geographic Sampling | 3 | 50-300 km | 10000 | ROC | 0.84858333 | 0.723 | 0.93 |
| Density-Weighted Geographic Sampling | 3 | 50-300 km | 10000 | TSS | 0.45266667 | 0.241 | 0.667 |
| Density-Weighted Geographic Sampling | 3 | 50-500km | 400 | ROC | 0.88866667 | 0.79 | 0.96 |
| Density-Weighted Geographic Sampling | 3 | 50-500km | 400 | TSS | 0.61925 | 0.381 | 0.868 |
| Density-Weighted Geographic Sampling | 3 | 50-500km | 800 | ROC | 0.88975 | 0.834 | 0.956 |
| Density-Weighted Geographic Sampling | 3 | 50-500km | 800 | TSS | 0.59225 | 0.237 | 0.859 |
| Density-Weighted Geographic Sampling | 3 | 50-500km | 2000 | ROC | 0.8885 | 0.813 | 0.943 |
| Density-Weighted Geographic Sampling | 3 | 50-500km | 2000 | TSS | 0.58408333 | 0.411 | 0.706 |
| Density-Weighted Geographic Sampling | 3 | 50-500km | 4000 | ROC | 0.86858333 | 0.781 | 0.945 |
| Density-Weighted Geographic Sampling | 3 | 50-500km | 4000 | TSS | 0.50483333 | 0.281 | 0.686 |
| Density-Weighted Geographic Sampling | 3 | 50-500km | 10000 | ROC | 0.84775 | 0.726 | 0.924 |
| Density-Weighted Geographic Sampling | 3 | 50-500km | 10000 | TSS | 0.46241667 | 0.25 | 0.663 |
| Density-Weighted Geographic Sampling | 5 | 0 | 400 | ROC | 0.83225 | 0.699 | 0.95 |
| Density-Weighted Geographic Sampling | 5 | 0 | 400 | TSS | 0.52033333 | 0.15 | 0.87 |
| Density-Weighted Geographic Sampling | 5 | 0 | 800 | ROC | 0.83508333 | 0.745 | 0.929 |
| Density-Weighted Geographic Sampling | 5 | 0 | 800 | TSS | 0.47041667 | 0.24 | 0.785 |
| Density-Weighted Geographic Sampling | 5 | 0 | 2000 | ROC | 0.84066667 | 0.783 | 0.921 |

| | | | | | | | |
|---|---|---|---|---|---|---|---|
| Density-Weighted Geographic Sampling | 5 | 0 | 2000 | TSS | 0.4215 | 0.212 | 0.61 |
| Density-Weighted Geographic Sampling | 5 | 0 | 4000 | ROC | 0.83091667 | 0.73 | 0.932 |
| Density-Weighted Geographic Sampling | 5 | 0 | 4000 | TSS | 0.38 | 0.204 | 0.522 |
| Density-Weighted Geographic Sampling | 5 | 0 | 10000 | ROC | 0.79875 | 0.677 | 0.913 |
| Density-Weighted Geographic Sampling | 5 | 0 | 10000 | TSS | 0.27691667 | 0.019 | 0.426 |
| Density-Weighted Geographic Sampling | 5 | 50-150 km | 400 | ROC | 0.87716667 | 0.803 | 0.944 |
| Density-Weighted Geographic Sampling | 5 | 50-150 km | 400 | TSS | 0.56858333 | 0.212 | 0.821 |
| Density-Weighted Geographic Sampling | 5 | 50-150 km | 800 | ROC | 0.86641667 | 0.741 | 0.956 |
| Density-Weighted Geographic Sampling | 5 | 50-150 km | 800 | TSS | 0.561 | 0.248 | 0.844 |
| Density-Weighted Geographic Sampling | 5 | 50-150 km | 2000 | ROC | 0.86108333 | 0.744 | 0.945 |
| Density-Weighted Geographic Sampling | 5 | 50-150 km | 2000 | TSS | 0.51333333 | 0.265 | 0.694 |
| Density-Weighted Geographic Sampling | 5 | 50-150 km | 4000 | ROC | 0.84408333 | 0.692 | 0.926 |
| Density-Weighted Geographic Sampling | 5 | 50-150 km | 4000 | TSS | 0.46091667 | 0.233 | 0.648 |
| Density-Weighted Geographic Sampling | 5 | 50-150 km | 10000 | ROC | 0.81266667 | 0.639 | 0.925 |
| Density-Weighted Geographic Sampling | 5 | 50-150 km | 10000 | TSS | 0.36516667 | 0.162 | 0.595 |
| Density-Weighted Geographic Sampling | 5 | 50-300 km | 400 | ROC | 0.88308333 | 0.813 | 0.958 |
| Density-Weighted Geographic Sampling | 5 | 50-300 km | 400 | TSS | 0.59275 | 0.322 | 0.88 |
| Density-Weighted Geographic Sampling | 5 | 50-300 km | 800 | ROC | 0.89941667 | 0.842 | 0.955 |
| Density-Weighted Geographic Sampling | 5 | 50-300 km | 800 | TSS | 0.63316667 | 0.376 | 0.86 |
| Density-Weighted Geographic Sampling | 5 | 50-300 km | 2000 | ROC | 0.89708333 | 0.827 | 0.945 |
| Density-Weighted Geographic Sampling | 5 | 50-300 km | 2000 | TSS | 0.57616667 | 0.311 | 0.807 |
| Density-Weighted Geographic Sampling | 5 | 50-300 km | 4000 | ROC | 0.87725 | 0.792 | 0.945 |
| Density-Weighted Geographic Sampling | 5 | 50-300 km | 4000 | TSS | 0.52275 | 0.302 | 0.675 |
| Density-Weighted Geographic Sampling | 5 | 50-300 km | 10000 | ROC | 0.84625 | 0.747 | 0.932 |
| Density-Weighted Geographic Sampling | 5 | 50-300 km | 10000 | TSS | 0.44833333 | 0.229 | 0.668 |
| Density-Weighted Geographic Sampling | 5 | 50-500km | 400 | ROC | 0.89908333 | 0.854 | 0.958 |
| Density-Weighted Geographic Sampling | 5 | 50-500km | 400 | TSS | 0.65983333 | 0.524 | 0.895 |
| Density-Weighted Geographic Sampling | 5 | 50-500km | 800 | ROC | 0.91216667 | 0.851 | 0.957 |

| | | | | | | | |
|---|---|---|---|---|---|---|---|
| Density-Weighted Geographic Sampling | 5 | 50-500km | 800 | TSS | 0.65058333 | 0.436 | 0.804 |
| Density-Weighted Geographic Sampling | 5 | 50-500km | 2000 | ROC | 0.9005 | 0.833 | 0.949 |
| Density-Weighted Geographic Sampling | 5 | 50-500km | 2000 | TSS | 0.59883333 | 0.384 | 0.771 |
| Density-Weighted Geographic Sampling | 5 | 50-500km | 4000 | ROC | 0.88141667 | 0.806 | 0.947 |
| Density-Weighted Geographic Sampling | 5 | 50-500km | 4000 | TSS | 0.55691667 | 0.299 | 0.692 |
| Density-Weighted Geographic Sampling | 5 | 50-500km | 10000 | ROC | 0.85833333 | 0.78 | 0.929 |
| Density-Weighted Geographic Sampling | 5 | 50-500km | 10000 | TSS | 0.487 | 0.296 | 0.68 |
| Geographic Sampling | - | 50-150 km | 400 | ROC | 0.86083333 | 0.775 | 0.952 |
| Geographic Sampling | - | 50-150 km | 400 | TSS | 0.4555 | 0.127 | 0.789 |
| Geographic Sampling | - | 50-300 km | 400 | ROC | 0.89233333 | 0.82 | 0.993 |
| Geographic Sampling | - | 50-300 km | 400 | TSS | 0.57475 | 0.277 | 0.833 |
| Geographic Sampling | - | 50-500km | 400 | ROC | 0.88041667 | 0.818 | 0.952 |
| Geographic Sampling | - | 50-500km | 400 | TSS | 0.59375 | 0.373 | 0.852 |
| Geographic Sampling | - | 50-150 km | 800 | ROC | 0.86725 | 0.79 | 0.955 |
| Geographic Sampling | - | 50-150 km | 800 | TSS | 0.558 | 0.283 | 0.798 |
| Geographic Sampling | - | 50-300 km | 800 | ROC | 0.8745 | 0.804 | 0.953 |
| Geographic Sampling | - | 50-300 km | 800 | TSS | 0.55208333 | 0.239 | 0.811 |
| Geographic Sampling | - | 50-500km | 800 | ROC | 0.8985 | 0.848 | 0.953 |
| Geographic Sampling | - | 50-500km | 800 | TSS | 0.57658333 | 0.239 | 0.817 |
| Geographic Sampling | - | 50-150 km | 2000 | ROC | 0.85041667 | 0.735 | 0.94 |
| Geographic Sampling | - | 50-150 km | 2000 | TSS | 0.45075 | 0.231 | 0.601 |
| Geographic Sampling | - | 50-300 km | 2000 | ROC | 0.88308333 | 0.817 | 0.965 |
| Geographic Sampling | - | 50-300 km | 2000 | TSS | 0.54525 | 0.192 | 0.795 |
| Geographic Sampling | - | 50-500km | 2000 | ROC | 0.89125 | 0.823 | 0.949 |
| Geographic Sampling | - | 50-500km | 2000 | TSS | 0.55983333 | 0.399 | 0.686 |
| Geographic Sampling | - | 50-150 km | 4000 | ROC | 0.83041667 | 0.719 | 0.926 |
| Geographic Sampling | - | 50-150 km | 4000 | TSS | 0.42766667 | 0.21 | 0.587 |
| Geographic Sampling | - | 50-300 km | 4000 | ROC | 0.8715 | 0.79 | 0.938 |

| | | | | | | | |
|---|---|---|---|---|---|---|---|
| Geographic Sampling | - | 50-300 km | 4000 | TSS | 0.50258333 | 0.273 | 0.66 |
| Geographic Sampling | - | 50-500km | 4000 | ROC | 0.87133333 | 0.814 | 0.935 |
| Geographic Sampling | - | 50-500km | 4000 | TSS | 0.50591667 | 0.34 | 0.683 |
| Geographic Sampling | - | 50-150 km | 10000 | ROC | 0.81025 | 0.681 | 0.91 |
| Geographic Sampling | - | 50-150 km | 10000 | TSS | 0.34691667 | 0.118 | 0.594 |
| Geographic Sampling | - | 50-300 km | 10000 | ROC | 0.84383333 | 0.757 | 0.914 |
| Geographic Sampling | - | 50-300 km | 10000 | TSS | 0.41708333 | 0.175 | 0.655 |
| Geographic Sampling | - | 50-500km | 10000 | ROC | 0.85983333 | 0.781 | 0.931 |
| Geographic Sampling | - | 50-500km | 10000 | TSS | 0.43666667 | 0.196 | 0.66 |
